# A 22-gene transcriptomic model indicating individual therapy durations in multidrug-resistant tuberculosis

**DOI:** 10.1101/2020.08.21.20177238

**Authors:** Jan Heyckendorf, Sebastian Marwitz, Maja Reimann, Korkut Avsar, Andrew DiNardo, Gunar Günther, Michael Hoelscher, Elmira Ibraim, Barbara Kalsdorf, Stefan H.E. Kaufmann, Irina Kontsevaya, Frank van Leth, Anna Maria Mandalakas, Florian Maurer, Marius Müller, Dörte Nitschkowski, Ioana D. Olaru, Cristina Popa, Andrea Rachow, Thierry Rolling, Jan Rybniker, Helmut J. F. Salzer, Patricia Sanchez-Carballo, Maren Schuhmann, Dagmar Schaub, Victor Spinu, Isabelle Suárez, Elena Terhalle, Markus Unnewehr, January Weiner, Torsten Goldmann, Christoph Lange

## Abstract

Emerging multidrug-resistant tuberculosis is a major global health challenge. The World Health Organization currently recommends treatment durations of 9–18 months or more for patients with multidrug-resistant tuberculosis. We identified and validated a host-RNA signature to serve as a biomarker for individualized therapy durations for patients with multidrug-resistant tuberculosis. Adult patients with pulmonary tuberculosis were prospectively enrolled into 5 independent cohorts in Germany and Romania. Clinical and microbiological data, and whole-blood for RNA transcriptomic analysis were collected at pre-defined timepoints throughout therapy. Treatment outcomes were ascertained one year after end-of-therapy. A whole-blood RNA therapy end model was developed in a multi-step process involving a machine-learning algorithm to identify hypothetical individual end-of-treatment timepoints. Fifty patients with drug-susceptible tuberculosis and 30 patients with multidrug-resistant tuberculosis were recruited in the German identification cohorts (DS- and MDR-GIC), 28 patients with drug-susceptible tuberculosis and 32 patients with multidrug-resistant tuberculosis in the German validation cohorts (DS- and MDR-GVC), and 52 patients with multidrug-resistant tuberculosis in the Romanian validation cohort (MDR-RVC). A 22-gene RNA model that defined cure-associated end-of-therapy timepoints was derived from the DS- and MDR-GIC data. The model accurately predicted clinical outcomes for patients in the DS-GVC (AUC=0.937 [95%CI:0.899–0.976]) and suggested that cure may be achieved with shorter treatment durations for tuberculosis patients in the MDR-GIC (mean reduction 218.0 days, 34.2%, p<0.001), the MDR-GVC (mean reduction 211.0 days, 32.9%, p<0.001), and the MDR-RVC (mean reduction of 161.0 days, 23.4%, p=0.001). Biomarker-guided management may substantially shorten the duration of therapy for many patients with multidrug-resistant tuberculosis.

**One Sentence Summary:** We identified and validated a transcriptome model based on a 22-gene signature to predict individual treatment durations for patients with multidrug-resistant tuberculosis.

## Introduction

Tuberculosis remains a major global health threat with emerging *Mycobacterium tuberculosis (M. tuberculosis*) drug-resistance being particularly worrisome (*1*). Multidrug-resistant tuberculosis (MDR-TB; defined by bacillary resistance against rifampicin and isoniazid) and extensively drug-resistant tuberculosis (XDR-TB; defined by multidrug-resistant tuberculosis plus resistance against at least one fluoroquinolone and one of the second-line injectable drugs amikacin, capreomycin and/or kanamycin) are associated with high treatment costs (*2*), frequently occurring adverse events (*3*), and discouragingly poor outcomes (*4*) despite prolonged treatment duration of 18 months or longer (*5, 6*). The World Health Organization (WHO) has endorsed a short-course multidrug-resistant tuberculosis regimen lasting 9–12 months (*7*) for patients with fluoroquinolone susceptible multidrug-resistant tuberculosis who also fulfil certain criteria. Nevertheless, the great majority of patients in several regions of the world, including Europe, are not eligible for the short-course regimen due to second-line *M. tuberculosis* drug-resistance (*8*).

The treatment duration needed to achieve cure is highly variable among individual patients and depends on the host’s immune status, the severity of disease, the pathogen’s virulence and drug-resistance status, as well as drug availability (*9, 10*). There is a growing interest and clinical need for a biosignature to guide individualized treatment duration (*11*); this is especially relevant for the treatment of patients with drug-resistant tuberculosis in order to reduce the rate of adverse events, cost, and to improve compliance (*9*).

Due to rapid changes in expression profiles following the initiation of anti-tuberculosis drug treatment, host genome-wide RNA expression holds promise as a surrogate marker for the duration of treatment required for an individual to achieve cure (*12*). RNA signatures that correlate with treatment response, and predict individual patient outcome including disease recurrence have been previously described in drug-susceptible tuberculosis patients (*13*).

We prospectively analyzed whole-blood RNA transcripts in patients from two identification cohorts. We aimed to develop a model indicating for cure in patients with drug-susceptible tuberculosis as a reference for which the host’s gene expression status under therapy was identified, and then applied this model to patients with multidrug-resistant tuberculosis to indicate individual end-of-therapy timepoints. Subsequently, this model was prospectively applied to three independent validation cohorts, one with drug-susceptible tuberculosis and two with multidrug-resistant tuberculosis.

## Results

All patients enrolled into these cohorts had culture confirmed pulmonary tuberculosis (**Table 1, Figure 1**) (*14, 15*). In detail, fifty patients with drug-susceptible tuberculosis were enrolled into the drug-susceptible German Identification Cohort (DS-GIC) and 30 MDR tuberculosis patients to the multidrug-resistant German Identification Cohort (MDR-GIC). Twenty-eight drug-susceptible tuberculosis patients were included in the drug-susceptible tuberculosis German Validation Cohort (DS-GVC), 32 MDR tuberculosis patients in the multidrug-resistant tuberculosis German Validation Cohort (MDR-GVC), and 52 MDR tuberculosis patients in the multidrug-resistant tuberculosis Romanian Validation Cohort (MDR-RVC). Patients were followed up one year after therapy end to assess for disease recurrence after therapy end. Clinical and mycobacterial data as well as transcriptomic data from samples taken longitudinally throughout therapy from the cohorts’ patients were available to conduct the analysis.

**Table 1:**
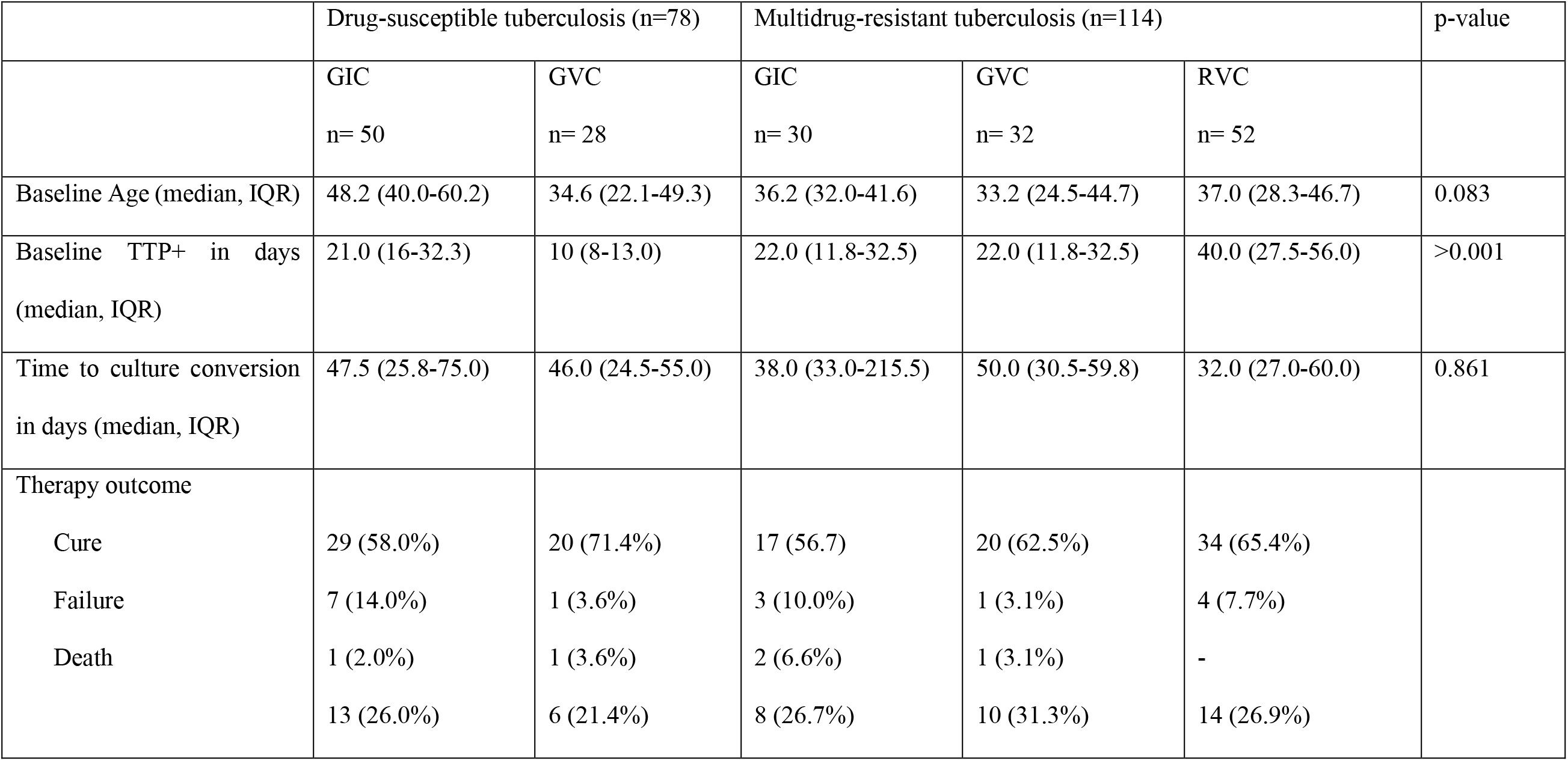
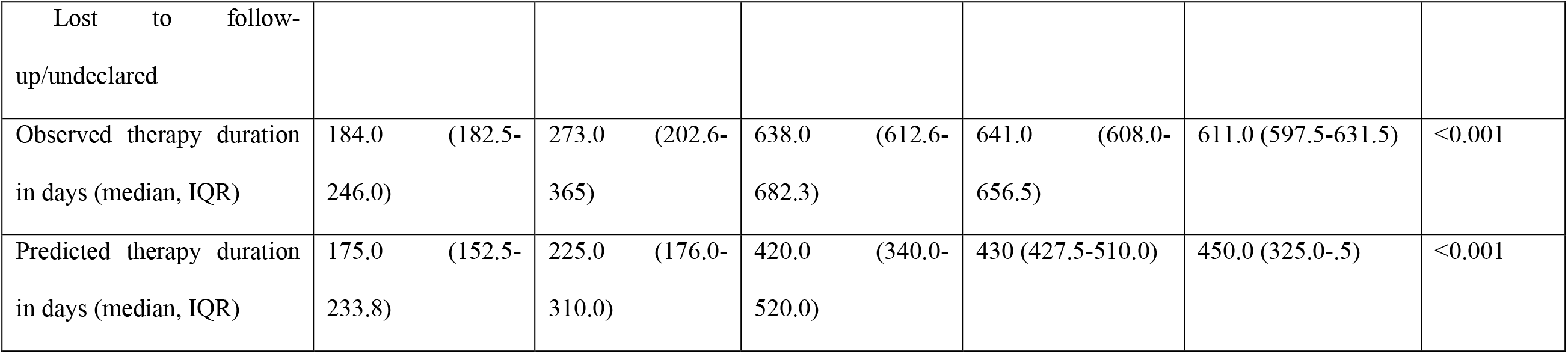
Clinical characteristics of tuberculosis patients including the observed and the predicted therapy durations in drug-susceptibleand multidrug-resistant tuberculosis patients from the German identification cohorts (GIC), the German validation cohorts (GVC), and the Romanian validation cohort (RVC). Therapy outcomes are derived following the TBNET criteria (*16*). TTP+ = time to culture positivity, IQR = interquartile range.

|  | Drug-susceptible tuberculosis (n=78) |  | Multidrug-resistant tuberculosis (n=114) |  |  | p-value |
| --- | --- | --- | --- | --- | --- | --- |
|  | GIC<br>n= 50 | GVC<br>n= 28 | GIC<br>n= 30 | GVC<br>n= 32 | RVC<br>n= 52 |  |
| Baseline Age (median, IQR) | 48.2 (40.0-60.2) | 34.6 (22.1-49.3) | 36.2 (32.0-41.6) | 33.2 (24.5-44.7) | 37.0 (28.3-46.7) | 0.083 |
| Baseline TTP+ in days<br>(median, IQR) | 21.0 (16-32.3) | 10 (8-13.0) | 22.0 (11.8-32.5) | 22.0 (11.8-32.5) | 40.0 (27.5-56.0) | >0.001 |
| Time to culture conversion<br>in days (median, IQR) | 47.5 (25.8-75.0) | 46.0 (24.5-55.0) | 38.0 (33.0-215.5) | 50.0 (30.5-59.8) | 32.0 (27.0-60.0) | 0.861 |
| Therapy outcome |  |  |  |  |  |  |
| Cure | 29 (58.0%) | 20 (71.4%) | 17 (56.7) | 20 (62.5%) | 34 (65.4%) |  |
| Failure | 7 (14.0%) | 1 (3.6%) | 3 (10.0%) | 1 (3.1%) | 4 (7.7%) |  |
| Death | 1 (2.0%) | 1 (3.6%) | 2 (6.6%) | 1 (3.1%) | - |  |
|  | 13 (26.0%) | 6 (21.4%) | 8 (26.7%) | 10 (31.3%) | 14 (26.9%) |  |
| Lost to follow-up/undeclared |  |  |  |  |  |  |
| Observed therapy duration in days (median, IQR) | 184.0 (182.5-246.0) | 273.0 (202.6-365) | 638.0 (612.6-682.3) | 641.0 (608.0-656.5) | 611.0 (597.5-631.5) | <0.001 |
| Predicted therapy duration in days (median, IQR) | 175.0 (152.5-233.8) | 225.0 (176.0-310.0) | 420.0 (340.0-520.0) | 430 (427.5-510.0) | 450.0 (325.0-.5) | <0.001 |

**Figure 1:**
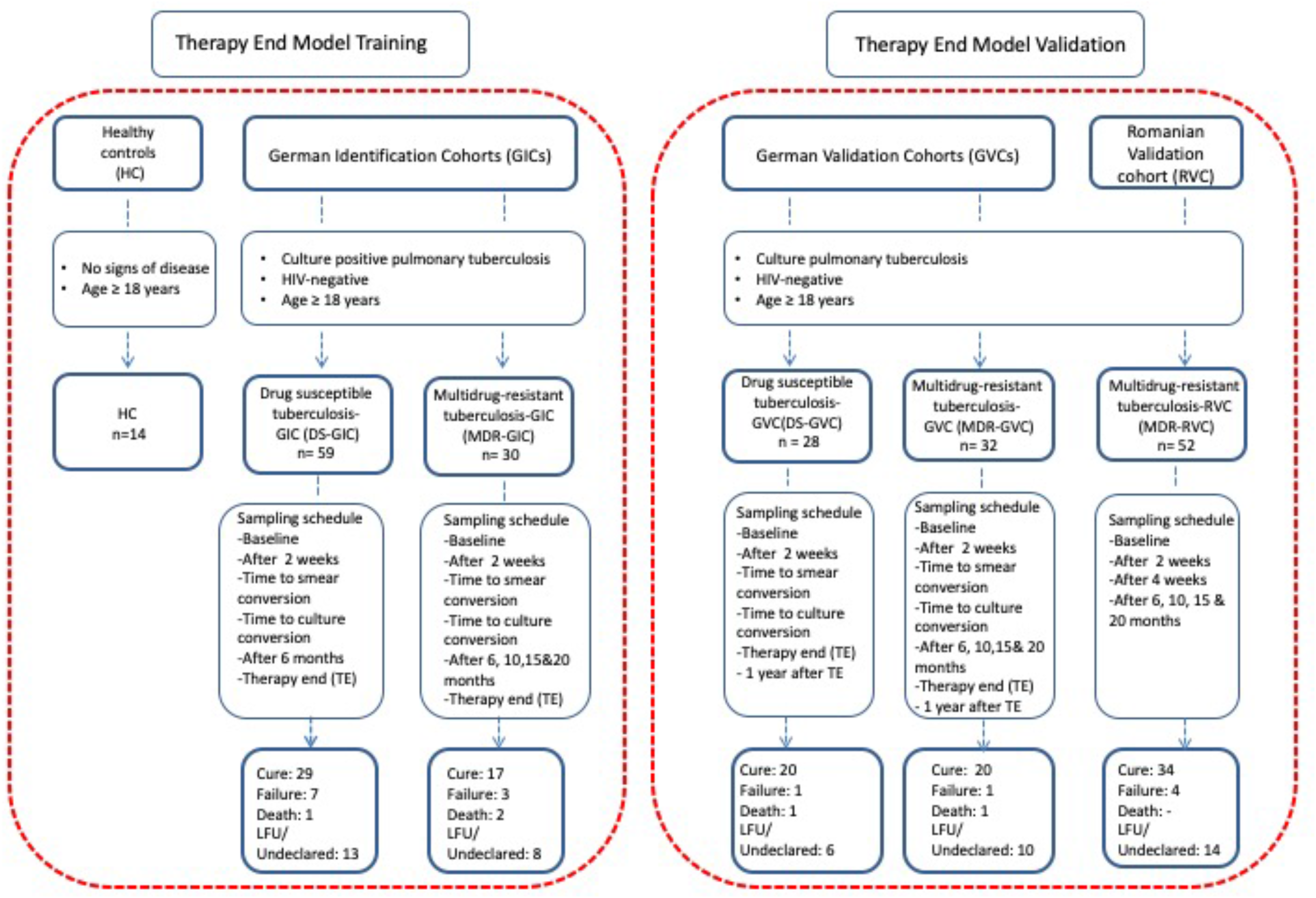
Cohorts of patients with drug-susceptible tuberculosis and multidrug-resistant tuberculosis. Flow chart showing recruitment for the model development of the five different cohorts. Patients with drug-susceptible (DS) and multidrug-resistant (MDR) tuberculosis were enrolled in the German Identification Cohorts (GICs), German Validation Cohorts (GVCs) and the Romanian Validation Cohort (RVC). Inclusion criteria were culture proven tuberculosis, sputum positivity at baseline, and age above 18 years at study inclusion. All patients were HIV negative. The blood sampling schedule for the different cohorts is depicted under the inclusion criteria. Therapy outcomes were assessed following the simplified TBNET definitions (*16*), which include a one year follow up after the end of therapy. Additionally, healthy controls (HC) were enrolled at the Research Center Borstel. The red dotted boxes indicate the use of the cohort’s data for either model identification (left) or validation (right).

All patients in the DS-GIC were infected with fully drug-susceptible strains of *M. tuberculosis*. In the DS-GVC, two patients (7.1%) had non-rifampicin polydrug-resistant tuberculosis with resistances to isoniazid and streptomycin, and to isoniazid and prothionamide, respectively, and one patient (3.1%) had isoniazid mono-resistant tuberculosis. Baseline time to sputum culture positivity (TTP+) in therapy naïve patients was not significantly different in the DS-GIC when compared to the DS-GVC (DS-GIC median 21 days, interquartile range [IQR]: 16.0–32.3 days vs. DS-GVC DS-TB median 10 days, IQR: 8.0–13.0 days; p=0.080). The median duration of therapy was 184 days (IQR: 182.5–246.0 days) in DS-GIC patients and 273 days (IQR: 202.6–365 days) in DS-GVC patients (p=0.038). MDR-GIC, MDR-GVC, and MDR-RVC patients were treated for a median duration of 638 days (IQR: 612.6–682.3 days), 641 days (IQR: 608.0–656.5 days), and 611 days (IQR 597.5–631.5 days), respectively (p=0.729). Patient outcomes can be found in **Figure 1 and Table 1**.

### Therapy end model conception

We conducted three steps to develop a unified model to calculate hypothetical end-of-therapy timepoints for DS-GVC patients (**Figure 2 and 3**). This was then further validated in independent data sets from the MDR-GIC, MDR-GVC, and MDR-RVC. This process also involved the use of clinical and mycobacterial data to evaluate the model’s plausibility. Therapy durations calculated by the model were hypothetical and retrospective, but were compared to the durations that were conducted in clinical reality.

**Figure 2:**
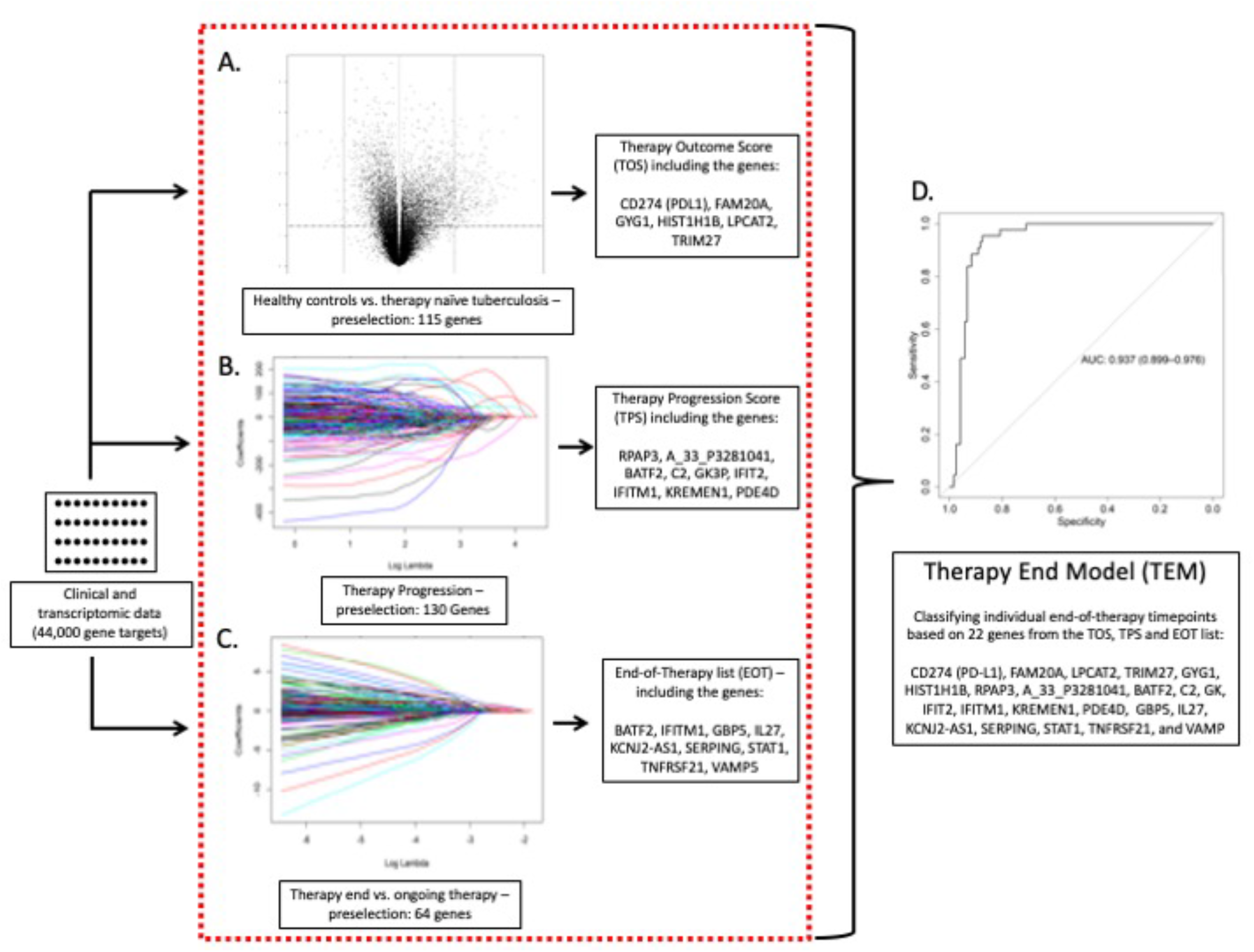
Multi-step development of the therapy end model for tuberculosis treatment. Simplified flow chart showing the multi-step approach of transcriptomic and clinical data analysis to develop the therapy end model (TEM) that identifies the optimal timepoint to stop anti-tuberculosis therapy. A. Development of *therapy outcome score (TOS)*. Showing the volcano plot representing differentially expressed genes in healthy controls vs. therapy naive drug-susceptible (DS-) and multidrugresistant (MDR) tuberculosis patients from the German identification cohorts (GICs). Genes that were significantly up- or downregulated (significant ≥2-fold change after Benjamini-Hochberg correction) form the basis for the TOS development. B. *Therapy progression score (TPS) development*. Depiction of penalizing regression coefficient adjustment (y-axis) and the explained deviation as a function of Log-ʎ (x-axis) for variable selection to identify genes that predict the remaining days of therapy that has been conducted in reality in all sample measurements from DS- and MDR-GIC tuberculosis patients. Each line represents one gene of interest and the genes shown in the plot were pre-selected by the initial Lasso regression step. The initial data selection was carried out on the entire data set with 44,000 gene targets. C. *End-of-therapy list (EOT list)*. Showing penalizing regression coefficient adjustment (y-axis) and the explained deviation as a function of Log-ʎ (x-axis) for variable selection to identify genes that classify between sample measurements in DS-GIC tuberculosis patients under therapy vs. timepoints at the end of relapse-free therapy in DS-GIC tuberculosis patients. Each line represents one gene of interest and the gene targets shown in the plot were pre-selected by the initial Lasso regression to reduce the number of genes of interest. D. *Therapy end model (TEM)*. Implementing the gene scores (TOS and TPS) and the EOT list into a machine learning algorithm model (Random Forest), a final simplified TEM for the calculation of end-of-therapy timepoints was developed via Generalized Linear Model (GLM). The initial TEM evaluation was carried out on data from DS-GIC tuberculosis patients. The ROC curve shows TEM’s classification accuracy in the independent data set of DS German validation cohort (GVC) tuberculosis patients (Area under the curve (AUC) 0.937, confidence interval (CI) 0.899–0.976)). TEM was further applied to patients with multidrug-resistant tuberculosis from the GIC, GVC, and from the Romanian validation cohort (MDR-RVC).

**Figure 3:**
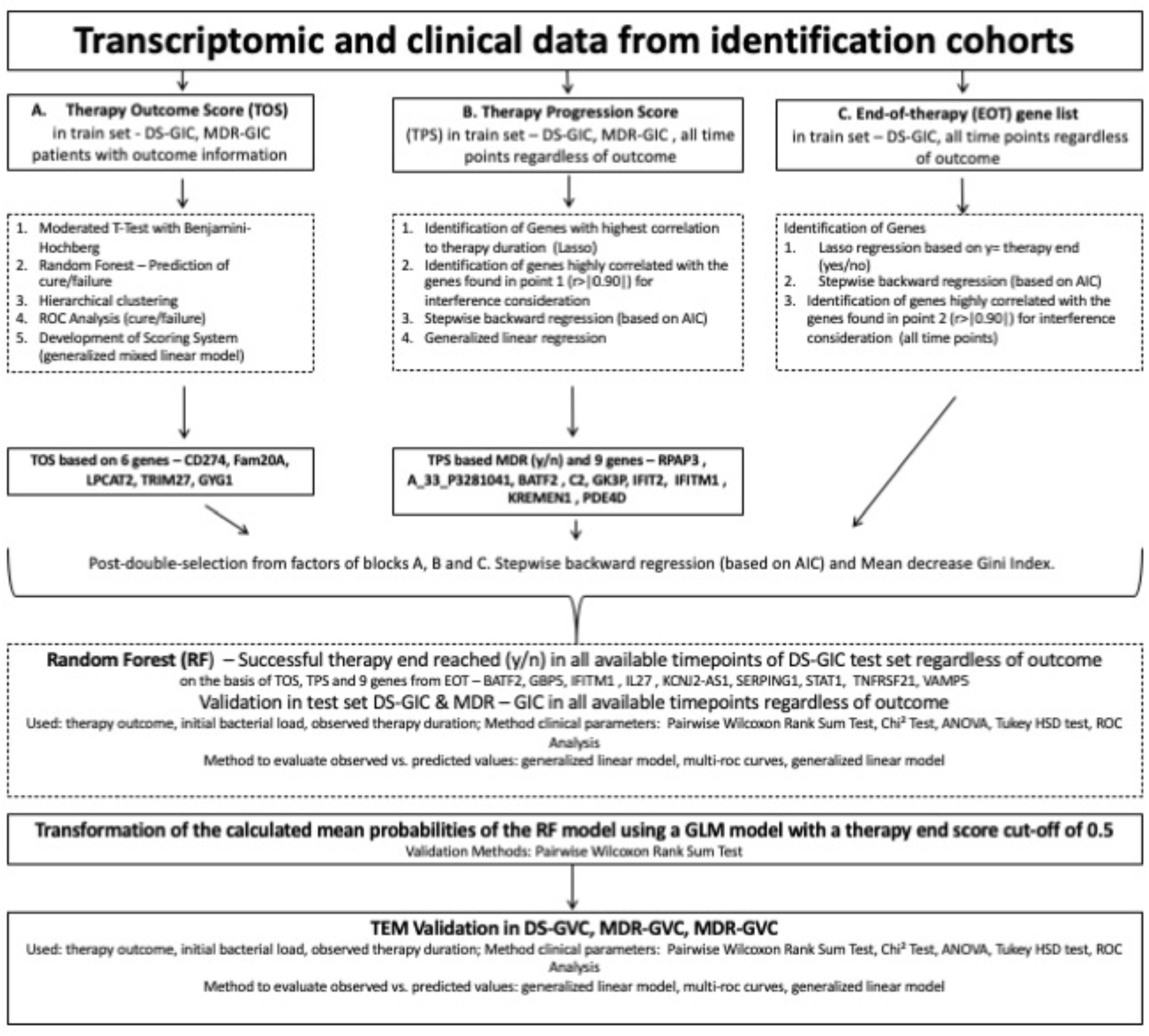
Development of therapy end model for tuberculosis treatment. Flow chart illustrating the therapy end model (TEM) development. A. Therapy outcome score (TOS). Identification of genes corresponding to therapy outcome (Cure/Failure/Death) according to TBNET criteria (*16*) by cross-validation in all drug susceptible (DS) and multidrug-resistant (MDR)-tuberculosis patients of the German Identification Cohorts (GICs). B. Therapy Progression Score (TPS). Identification of genes that correlate with the remaining therapy by cross-validation in DS- and MDR-tuberculosis patients of the GIC via Lasso regression. C. End-of-therapy (EOT) list. Identification of genes that are differently expressed previously to and at successful completion of therapy. The combination of TOS, TPS and the EOT list were included into the TEM in cross-validated DSGIC-patients. This model was then applied to independent data sets from DS-TB patients of the German validation cohort (GVC), and subsequently in patients from the MDR-GIC, the MDR-GVC, and the MDR-Romanian Validation Cohort (RVC).

The workflow that was followed to analyze transcriptome data is described in **Figure 2 and Figure 3**. In brief, transcriptome and clinical data from DS- and MDR-GIC were analyzed to build a model to predict therapy outcomes and to assess the progress of therapy. The resulting scores and a list of genes that characterized the end of successful therapy best in DS-GIC patients were integrated into a final model to determine individual end-of-therapy timepoints. This algorithm was then validated on transcriptome data of the independent GVCs and the RVC (See **Figure 1 and 3**). This validation process also involved the validation of the model’s results with clinical data such as times of culture and smear conversion (TCC, TSC), time of mycobacterial culture positivity (TTP+), the radiological extent of disease as well as presence of cavities in chest X-ray imaging. The following passages will address the development of the different scores and the gene list contributing to a final therapy end model that is able to identify end-of-therapy timepoints for tuberculosis patients.

#### Therapy outcome score (step 1)

A score correlating with therapy outcome at any given sampling timepoint was developed by identifying genes that were significantly up- or downregulated in therapy-naïve DS-GIC and MDR-GIC patients compared to healthy controls (HCs) in a moderated T-test with a fold-change cut-off of ≥2 and a Benjamini-Hochberg multiple testing correction of p≤0.01 (**Figure 2, Table 2, Supplementary File**). Next, this gene list was further analyzed to identify genes with the potential to predict patient therapy outcomes according to TBNET definitions (*16*) (outcome groups: cure, failure, death; For outcome definitions see **Table 3**; for further details regarding the analysis see **Table 2**, **Figure 2 and 3**) by using hierarchical clustering and cross-validated Random Forest model fitting in DS-GIC and MDR-GIC patients with available final therapy outcome data. The expression levels of six genes - CD274 (PDL1), FAM20A, GYG1, LPCAT2, HIST1H1B, and TRIM27 - were identified as an ideal model to distinguish between patients experiencing cure, failure or death following the TBNET outcome criteria (*16*). This procedure included the numerical translation of the outcome cure and non-favorable outcomes such as failure and death (dependent variable), which were described by the independent variables (expression of the six genes as described above) in a Generalized Linear Model (GLM) to form the therapy outcome score (see **Table 2, Figure 3**).

**Table 2:**
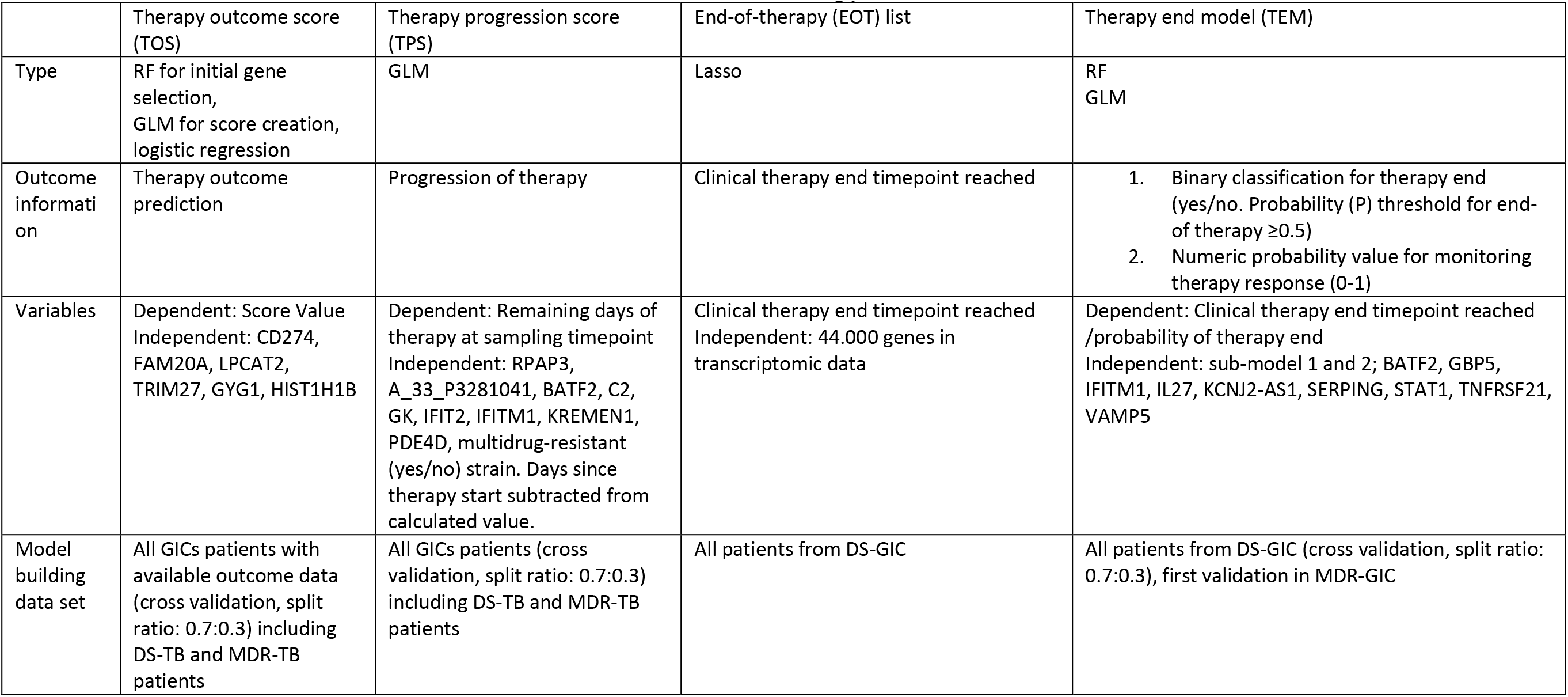
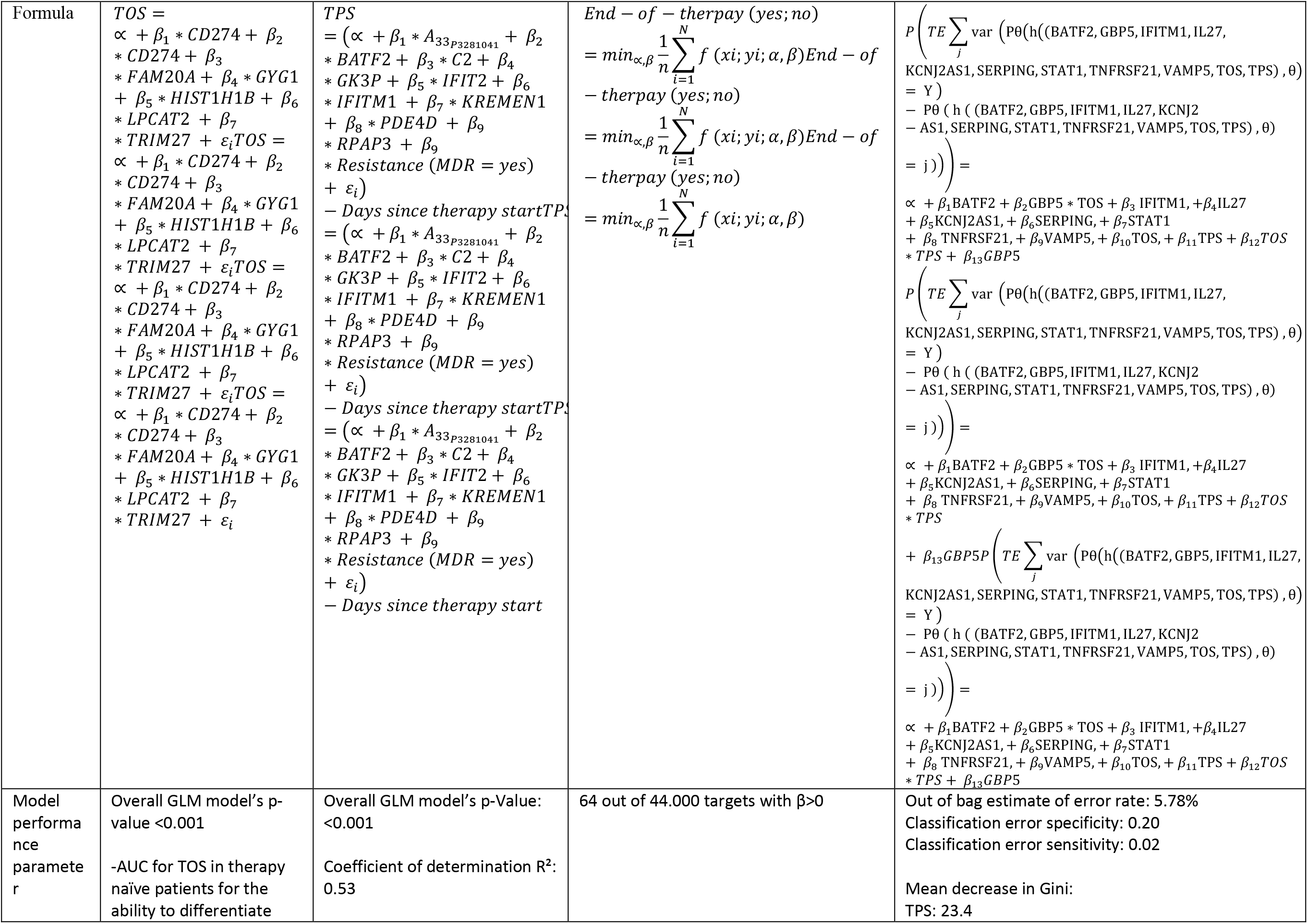
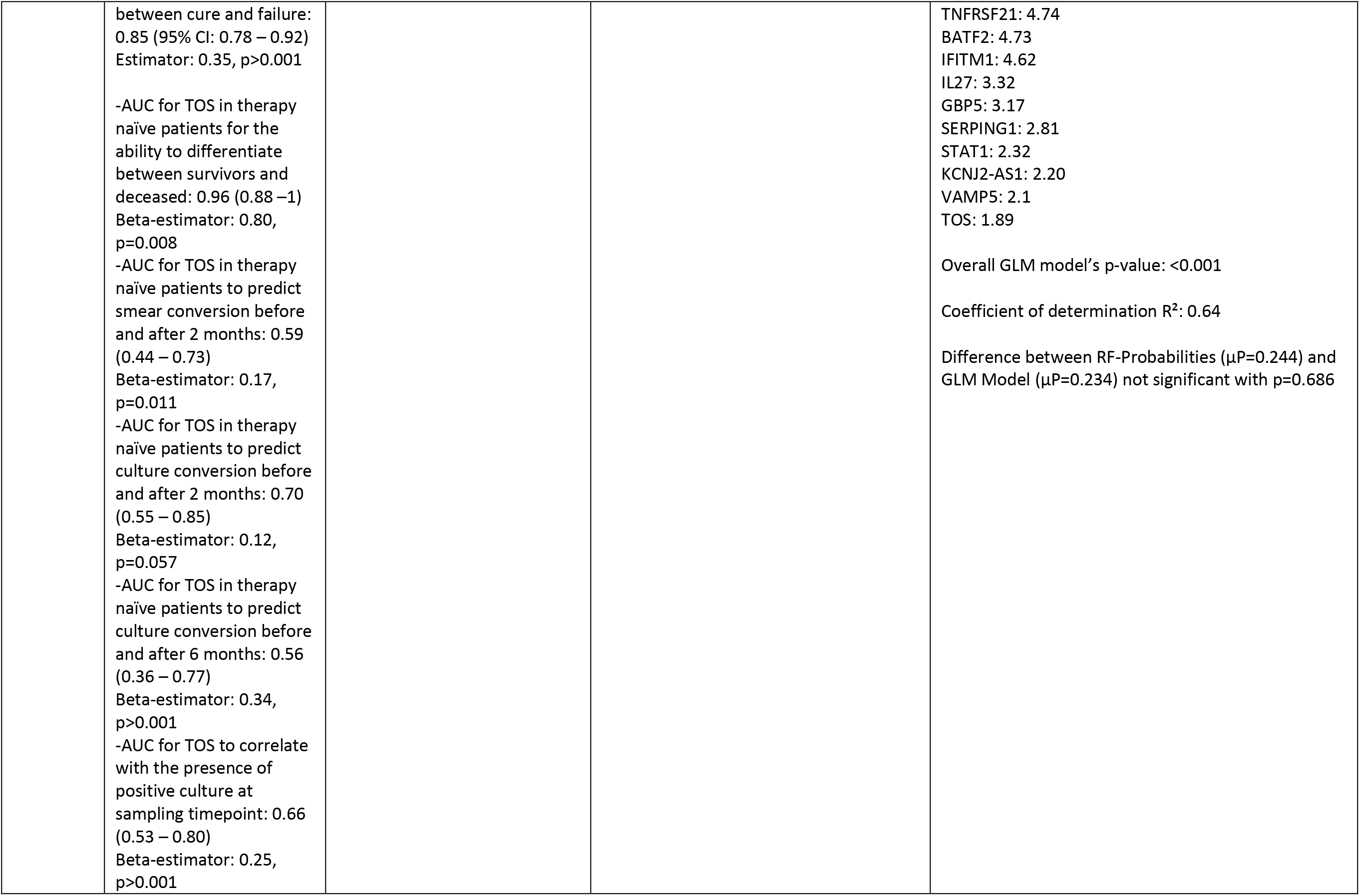
Model parameters of three steps leading to the end-of-therapy model for patients with tuberculosis. TOS= therapy outcome score, TPS = Therapy Progression Score, RF= Random Forest, DS= drug-susceptible, MDR= multidrug-resistant, TB= tuberculosis, GIC= German validation cohort, AUC= area under the curve, TE=Therapy End, GLM = Generalized linear model.

Further validation of this finding was performed by using the patients’ culture conversion status, the drug-resistance status, and the radiographic extent of disease at baseline to re-assure the plausibility of the therapy outcome score. Kruskal-Wallis test showed highly significant differences of the therapy outcome score between outcome groups at baseline in the GICs (p<0.001). MDR- and DS-GIC patients with prolonged times of culture conversion (2-month culture conversion status) also showed higher baseline score values (median: 3.4, IQR: 1.6–5.0) when compared to those with early culture conversion (median: 1.8, IQR: 0.5–3.4, p=0.111). Therapy naïve MDR- and DS-GIC patients experiencing cure had a median score of 0.86 (IQR: - 0.37–2.3) while patients with therapy failure exhibited a median score of 1.3 (IQR: 0.34–4.61) and deceased patients a value of 5.83 (IQR: 5.08–10.04; **Figure 4**). The performance of this outcome score to predict therapy outcome in therapy naïve patients was 0.85 (95% CI 0.78–0.92).

**Figure 4:**
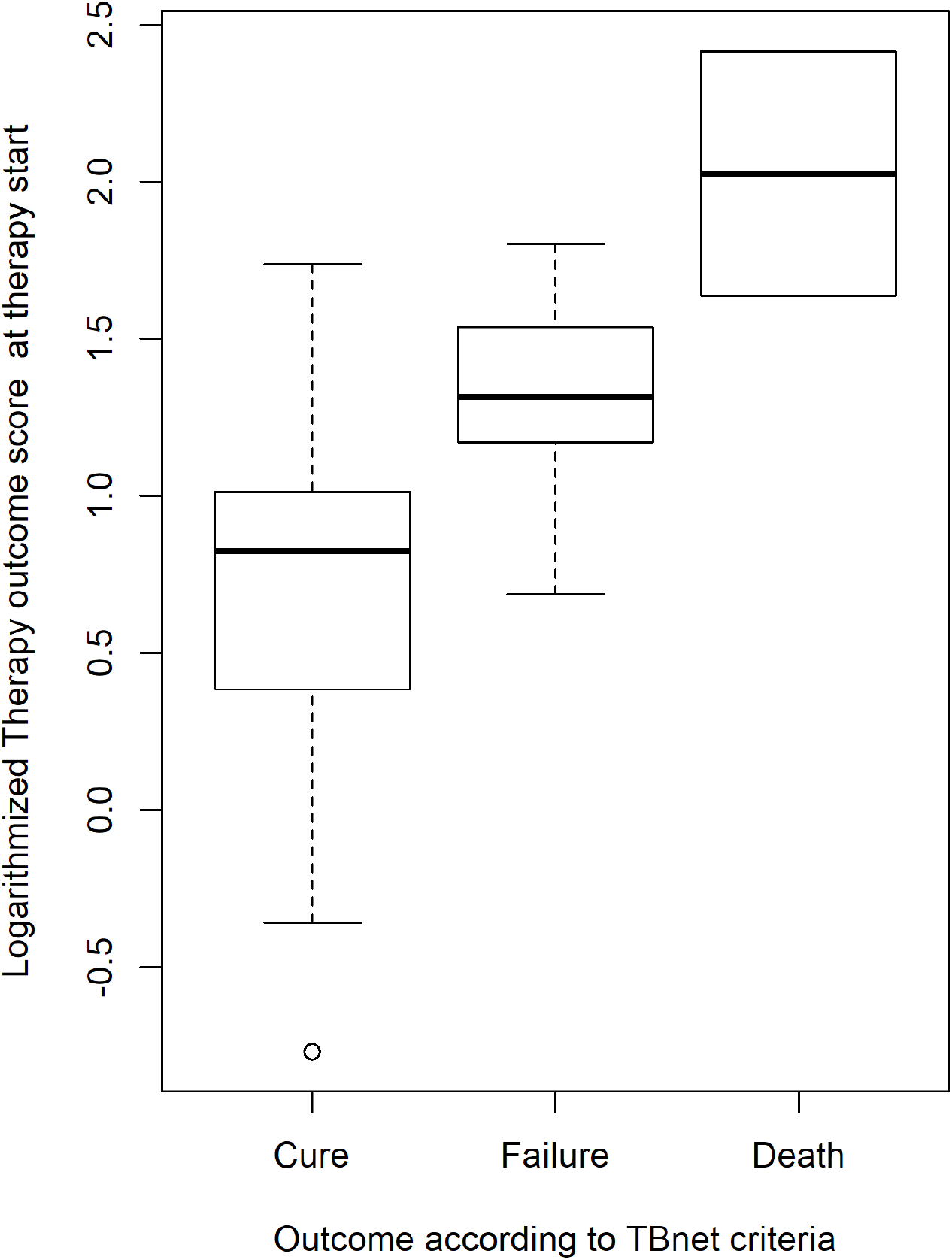
Therapy outcome score values in therapy-naïve patients with drug- susceptible tuberculosis and multidrug-resistant tuberculosis. Boxplot graph showing the comparison of logarithmic therapy outcome score (TOS) values in therapy naïve drug susceptible and multidrug-resistant tuberculosis patients from the German identification cohorts (GIC) with regard to their therapy outcome (cure, failure, or death according to the TBNET-criteria) (*16*).

#### Therapy progression score (Step 2)

To identify genes that correlate with the proximity (in days) to the clinical end-of-therapy for any given sample timepoint, we performed a pre-selection from the total transcriptomic data set of DS- and MDR-GICs by using penalizing regression model (Lasso; **Figure 2 and 3, Table 2**). Using this list of genes (**Supplementary File**), variable reduction steps were conducted by applying Bonferroni correction and stepwise Akaike information criterion (AIC; both directions with 1.000 steps) to focus on genes that highly correlated with the progress of therapy leading to a GLM-model consisting of nine genes (RPAP3, A_33_P3281041, BATF2, C2, GK3P, IFIT2, IFITM1, KREMEN1, PDE4D) and the patients’ drug resistance status (DS-TB vs. MDR-TB, binary variable). The time under therapy (in days) is subtracted from the GLM result to calculate the therapy progression score for each patient at any given sample timepoint, which correlated with the days of remaining therapy that was conducted in reality (r=0.59 (p<0.001); see **Table 2, Figure 3**).

#### End-of-therapy list (Step 3)

To further characterize the correlations with successful end-of-therapy for tuberculosis patients on the transcriptional level, we identified genes that were differentially expressed in patients under anti-tuberculosis therapy when compared to patients at the end of successful therapy (cure; see **Table 2, Figure 1-3**) (*16*). For this purpose, Lasso regression was performed throughout the cross-validated total gene expression data set of DS-GIC data to identify genes corresponding to sampling timepoints that would correspond to cure (**Table 2; Figure 1-3**). The genes identified by this process (**Supplementary File**) went through a further variable reduction process (Bonferroni correction and stepwise AIC method in both directions with 1.000 steps) resulting in a list of nine relevant gene targets from the end-of-therapy list (BATF2, GBP5, IFITM1, IL27, KCNJ2-AS1, SERPING1, STAT1, TNFRSF21, VAMP5). Two of these targets were also part of the therapy progression score (BATF2 and IFITM1).

### Therapy end model

The above described three steps were used as basis to develop the final therapy end model (**Table 2, Figure 1-3**). In summary, first, a set of six genes (CD274 [PDL-1], FAM20A, HIST1H1B, LPCAT2, GYG1, TRIM27) was identified with significant up- or downregulation in therapy-naïve DS-GIC and MDR-GIC patients compared to HC that were translated into the therapy outcome score. In the second step, a set of 9 gene targets (RPAP3, A_33_P3281041, BATF2, C2, GK, IFIT2, IFITM1, KREMEN1, PDE4D) was identified to determine genes that correlated with the remaining time of therapy in patients of the DS-GIC and MDR-GIC in longitudinal timepoints, which were implemented into the therapy progression score. The third step resulted in a list of target genes that characterized the transcriptional difference between patients under therapy and at the end-of-therapy, which included 9 genes (again the targets BATF2 and IFITM1, and additionally GBP5, IL27, KCNJ2-AS1, SERPING, STAT1, TNFRSF21, VAMP5; see **Table 2**, **Figure 3**).

The numerical values from the treatment outcome score, the therapy progress score and the end-of therapy list were then integrated into the therapy end model, which, after further reduction of variables using cross-validation and Mean Decrease Gini Index, resulted in a Random Forest (RF) model to classify for relapse-free cure end-of-therapy timepoints (**Table 2, Figure 1-3**). 5000-fold RF iterations resulted in average probabilities for the ideal end-of-therapy timepoint in individual DS-GIC patients at any given sample timepoint and validated in MDR-GIC. The threshold value for the successful end of therapy was set at P≥0.5 (standard RF classification threshold). The model’s results were then further validated and simplified by applying a Generalized Linear Model (GLM) to calculate the end-of-therapy score again with a cut-off of ≥0.5. The GLM was developed using the same variables as for the RF model, which yielded the same probability results and then served as final therapy end model (**Table 2**).

The final therapy end model consisted of a total of 22 gene targets (CD274 [PD-L1], FAM20A, LPCAT2, TRIM27, GYG1, HIST1H1B, RPAP3, A_33_P3281041, BATF2, C2, GK, IFIT2, IFITM1, KREMEN1, PDE4D, GBP5, IL27, KCNJ2-AS1, SERPING, STAT1, TNFRSF21, VAMP5) to calculate end-of-therapy scores at different timepoints (**Table 2, Figure 1-3**). The therapy end model identified end-of-therapy timepoints with high accuracy in DS-GVC patients (area under the curve [AUC] 0.937; 95% confidence interval [CI]:0.899–0.976; **Figure 5**). The model was applied to MDR-GIC, and to patients from the independent DS- and MDR-tuberculosis German Validation Cohort (GVC), and patients from the MDR-tuberculosis Romanian Validation Cohort (RVC) to calculate hypothetical therapy durations. **Figure 6A-C** shows the end-of-therapy probabilities of the different cohorts as a function of time under therapy. Each measurement resembles an independent end-of-therapy calculation for a tuberculosis patient under therapy. All calculation results above the cut-off ≥0.5 indicate for hypothetical end-of-therapy timepoints with cure as final treatment outcome.

**Figure 5:**
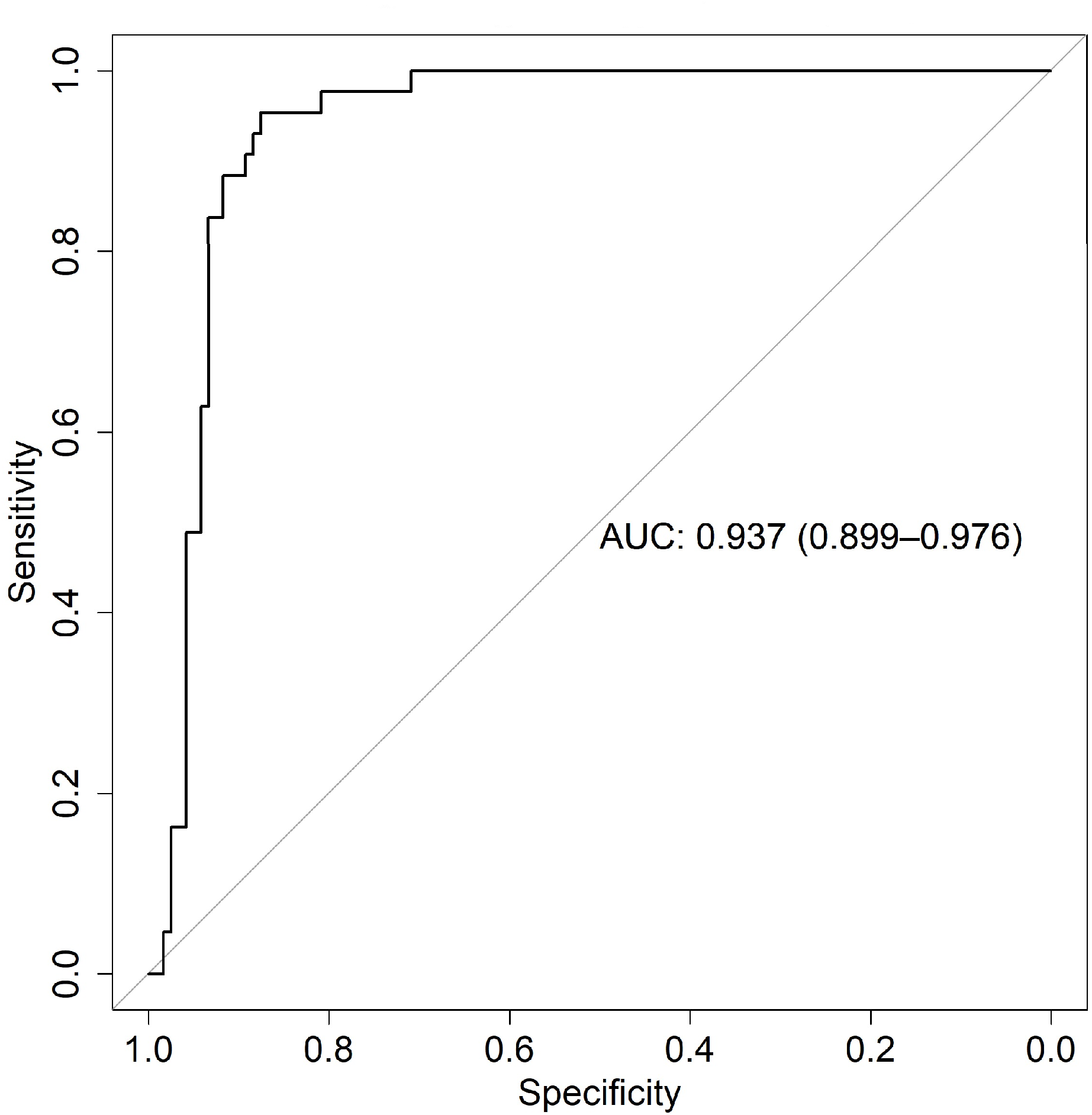
Receiver operating characteristic curve for the therapy end model classification in drug-susceptible tuberculosis patients from the German Validation Cohort. Receiver operating characteristic curve (ROC) analysis to evaluate the therapy end model’s performance to classify calculated drug susceptible (DS) German Validation Cohort patients (GVC) end-of-therapy timepoints when compared to clinical therapy end timepoints (area under the curve (AUC)=0.937 (95% confidence interval (CI) 0.899– 0.976).

**Figure 6:**
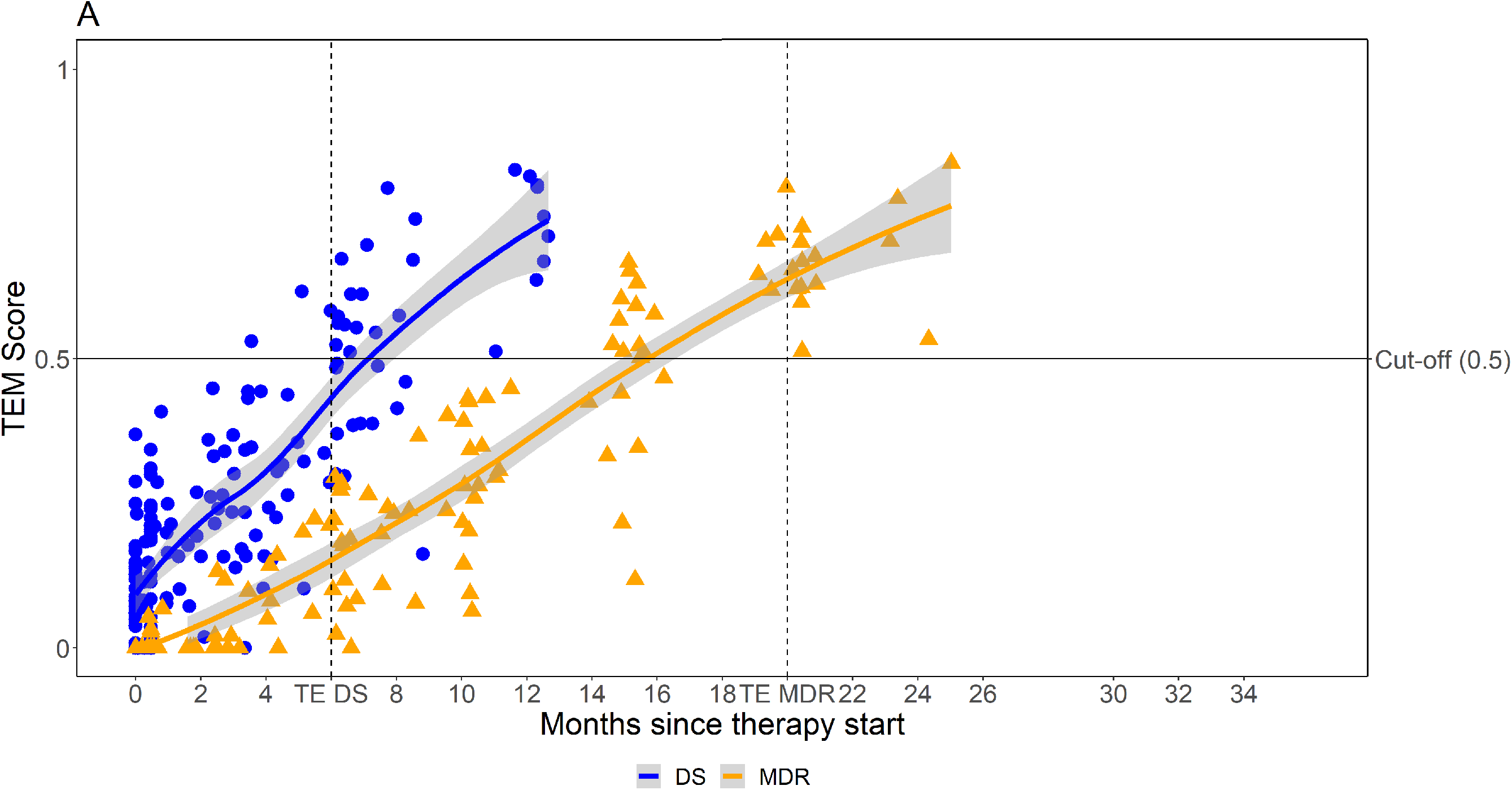
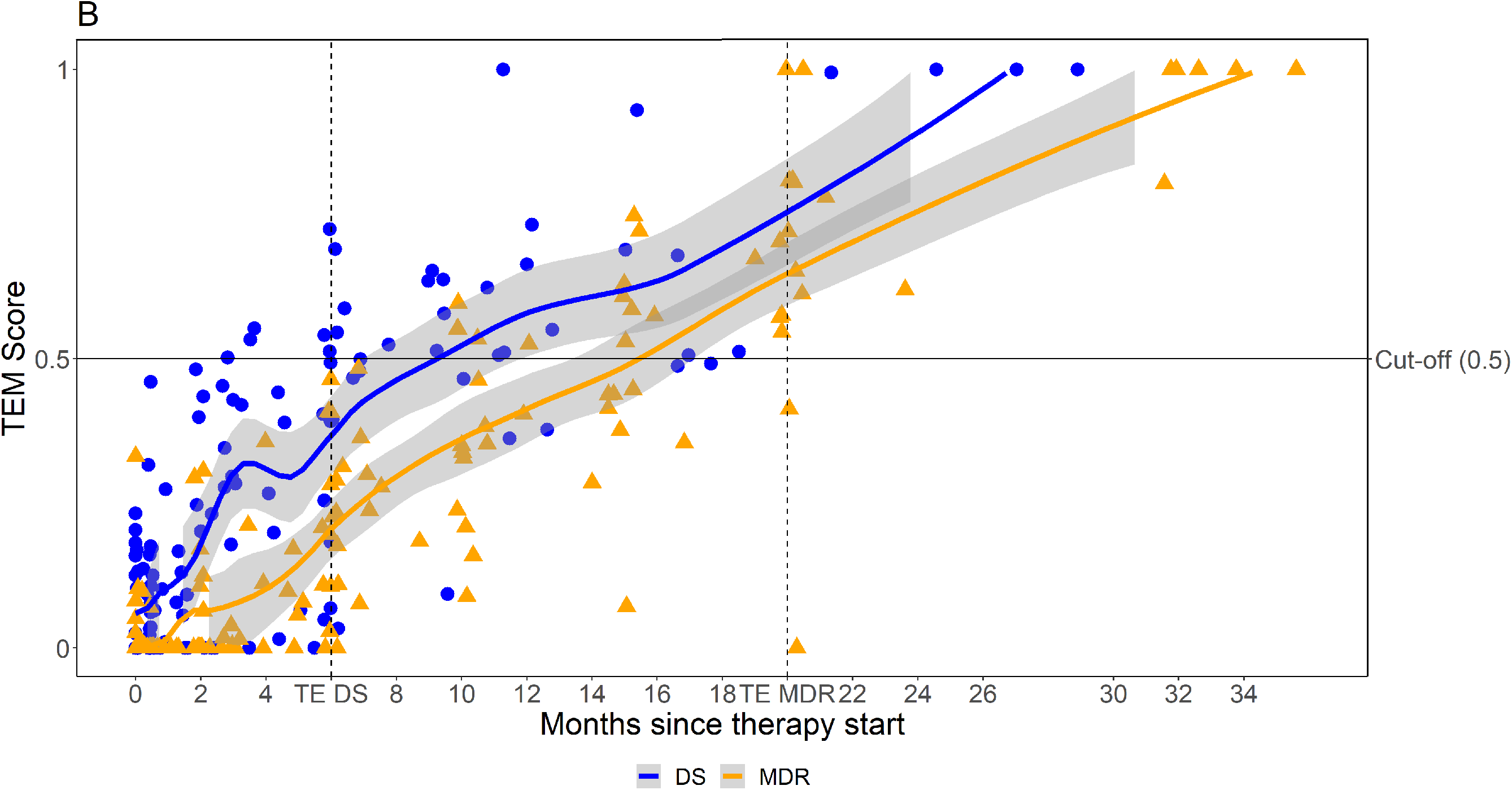
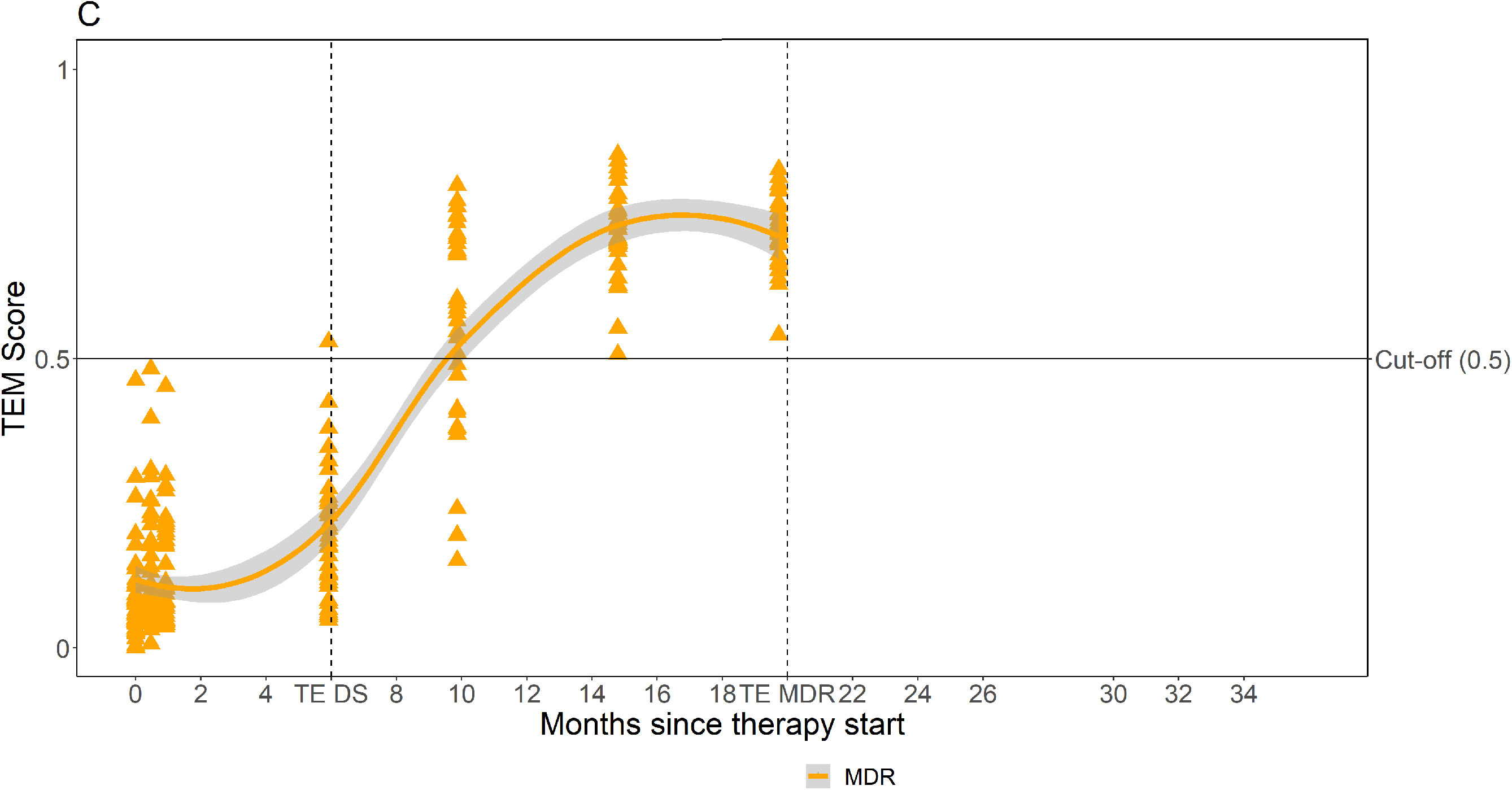
Therapy end model scores over the course of therapy for the five cohorts of patients with tuberculosis. Scores for end-of-therapy by the therapy end model (TEM) over the time of anti-tuberculosis treatment for the five cohorts of drug-susceptible (DS) tuberculosis and multidrug-resistant (MDR) tuberculosis patients of German Identification Cohort (GIC), German Validation Cohort (GVC) and Romanian Validation Cohort (RVC) following the therapy end model. Y-axis: TEM scores for end-of-therapy, horizontal line: probability threshold (P≥0.5) for relapse-free end-of-therapy; X-axis: time under treatment (months), first vertical dotted line indicates 6 months of therapy, the common timepoint of therapy end (TE) in drug-susceptible tuberculosis, second vertical dotted line indicates the usual timepoint for TE in multidrug-resistant tuberculosis after 20 months of therapy. **Figure 6A**: TEM scores in DS-GIC and MDR-GIC over time (blue: DS-GIC, orange: MDR-GIC) **Figure 6B**: TEM probabilities in DS-GVC and MDRGVC patients over time (blue DS-GVC, orange: MDR-GVC) **Figure 6C**: TEM scores in MDR-RVC patients over time (orange).

The proportion of patients who reached the model’s threshold for the calculated end-of-therapy at the end of clinical anti-tuberculosis treatment was 100% in the DS-GIC and 97.4% in the DS-GVC. Patients who did not reach the threshold indicating a relapse-free end-of-therapy at month 6 showed an increased time to *M. tuberculosis* sputum culture conversion when compared to those who did (median of 68 days, IQR: 50.0–126.0 days vs. median of 46.0 days, IQR: 30.0–63.0 days; p=0.041). None of the patients in the MDR-GIC and MDR-GVC and only one patient (1.9%; culture conversion within 2 weeks) in the MDR-RVC reached the threshold for cure at 6 months. Following 15 months of therapy, the overall proportions of multidrug-resistant tuberculosis patients having reached cure according to the model were 84.6% in the MDR-GIC, 40% in the MDR-GVC and 88.5% in the MDR-RVC.

When the therapy end model probabilities for therapy end were stratified for drug-resistance status in pooled data from the different cohorts, they showed low probabilities for therapy end at baseline, after 2 weeks of therapy, at smear and culture conversion, but probabilities above the threshold at clinical therapy end timepoints (**Figure 7A and B**). The therapy end model probabilities for end-of-therapy were also compared between patients with drug-susceptible tuberculosis and with multidrug-resistant tuberculosis at relevant bacteriologically defined endpoints such as the individual time of sputum culture and smear microscopy conversion (**Figure 8A-F**). End-of-therapy probabilities were well below the threshold for both drug-susceptible tuberculosis and multidrug-resistant tuberculosis at these timepoints, but probability values were significantly lower for patients with multidrug-resistant tuberculosis when compared to drug-susceptible tuberculosis patients in the GICs (median probability at smear conversion: DS-GIC P=0.21 vs. MDR-GIC P=0.06, p=0.038; median probability at culture conversion: DS-GIC P=0.29 vs. MDR-GIC P=0.04, p=0.007) and the GVCs (median probability at smear conversion: DS-GVC P=0.09 vs. MDR-GVC P=0.01, p=0.040; median probability at culture conversion: DS-GVC P=0.29 vs. MDR-GVC P=0.04, p=0.007). Of note, no patient with positive sputum culture results had a therapy end model calculation result for the end-of-therapy.

**Figure 7:**
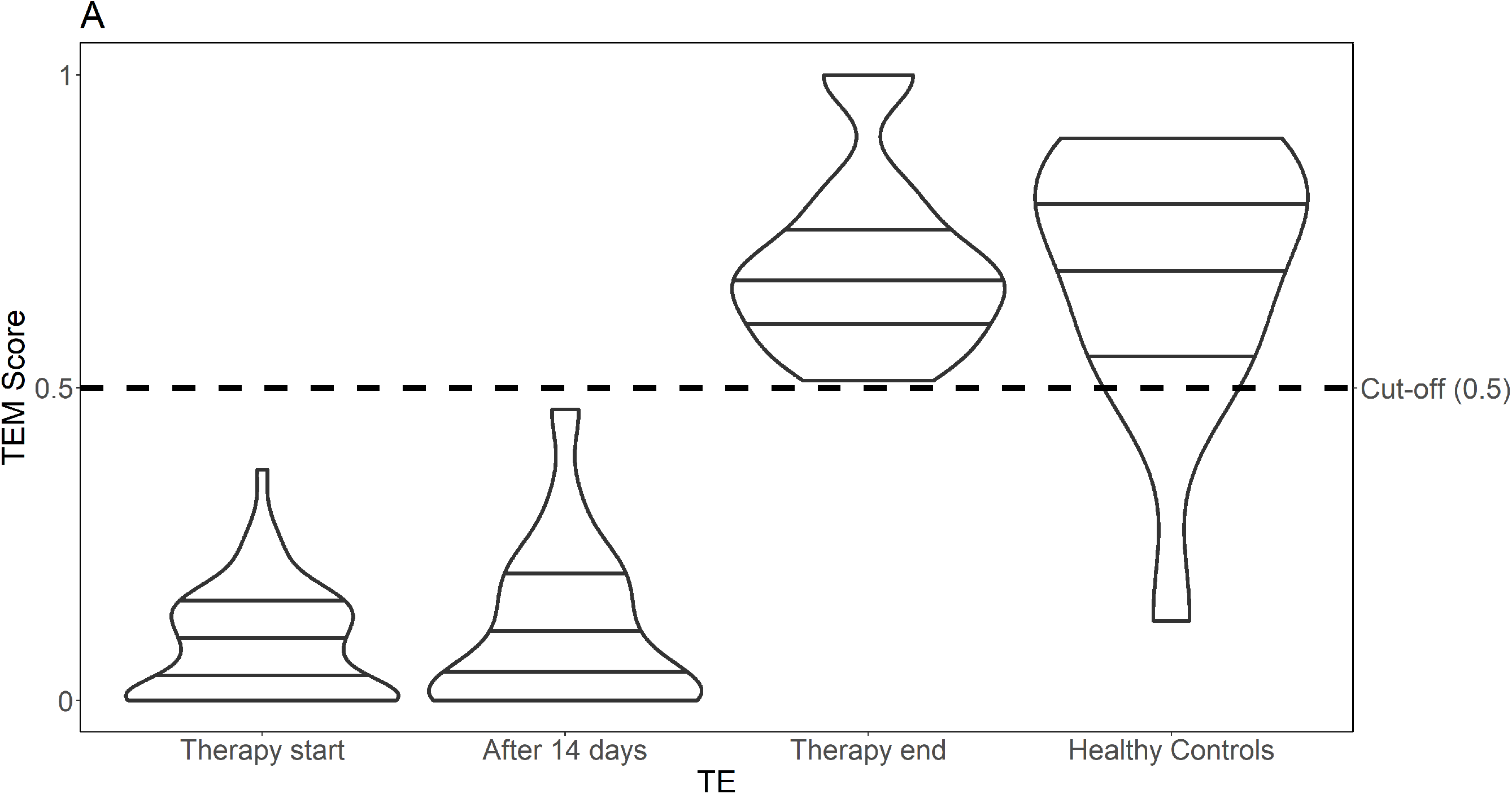
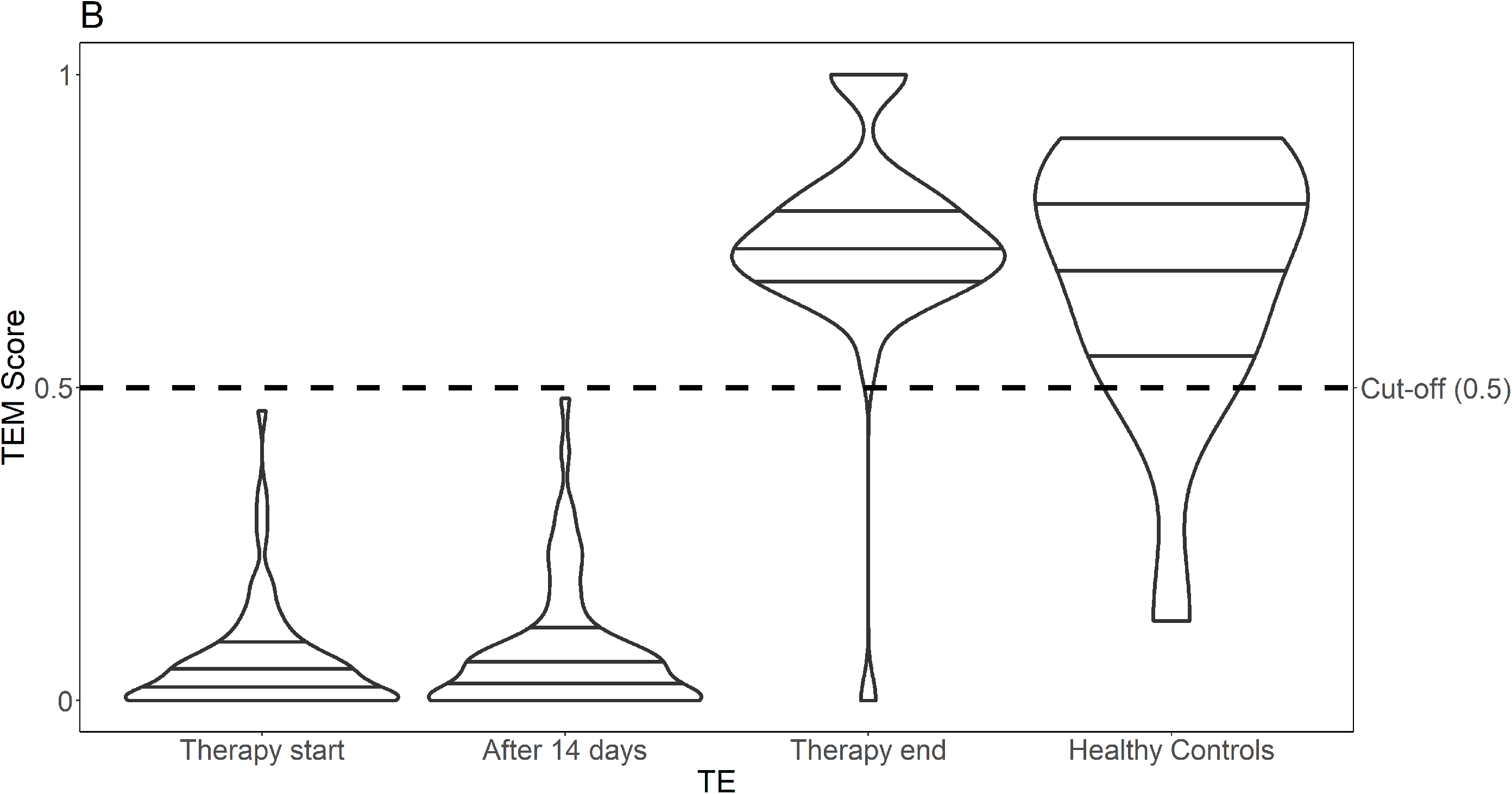
Therapy end model scores in patients with drug-susceptible tuberculosis and multidrug-resistant tuberculosis compared to healthy controls at relevant timepoints. Violin plots comparing calculated end-of-therapy scores of the therapy end model (TEM) for healthy controls, multidrug-resistant (MDR)- and drug-susceptible (DS) tuberculosis patients at relevant timepoints. Y-axis: TEM scores, dotted horizontal line: score threshold (Score (P)≥0.5) for relapse-free end-of-therapy; X-axis showing TEM scores at therapy start, after 14 days of therapy, at therapy end, and for healthy controls. **Figure 7A:** TEM scores for patients with DS tuberculosis at therapy start, after 14 days of therapy, at therapy end, and for healthy controls, **Figure 7B:** TEM scores for patients with MDR tuberculosis at therapy start, after 14 days of therapy, at therapy end, and for healthy controls.

**Figure 8:**
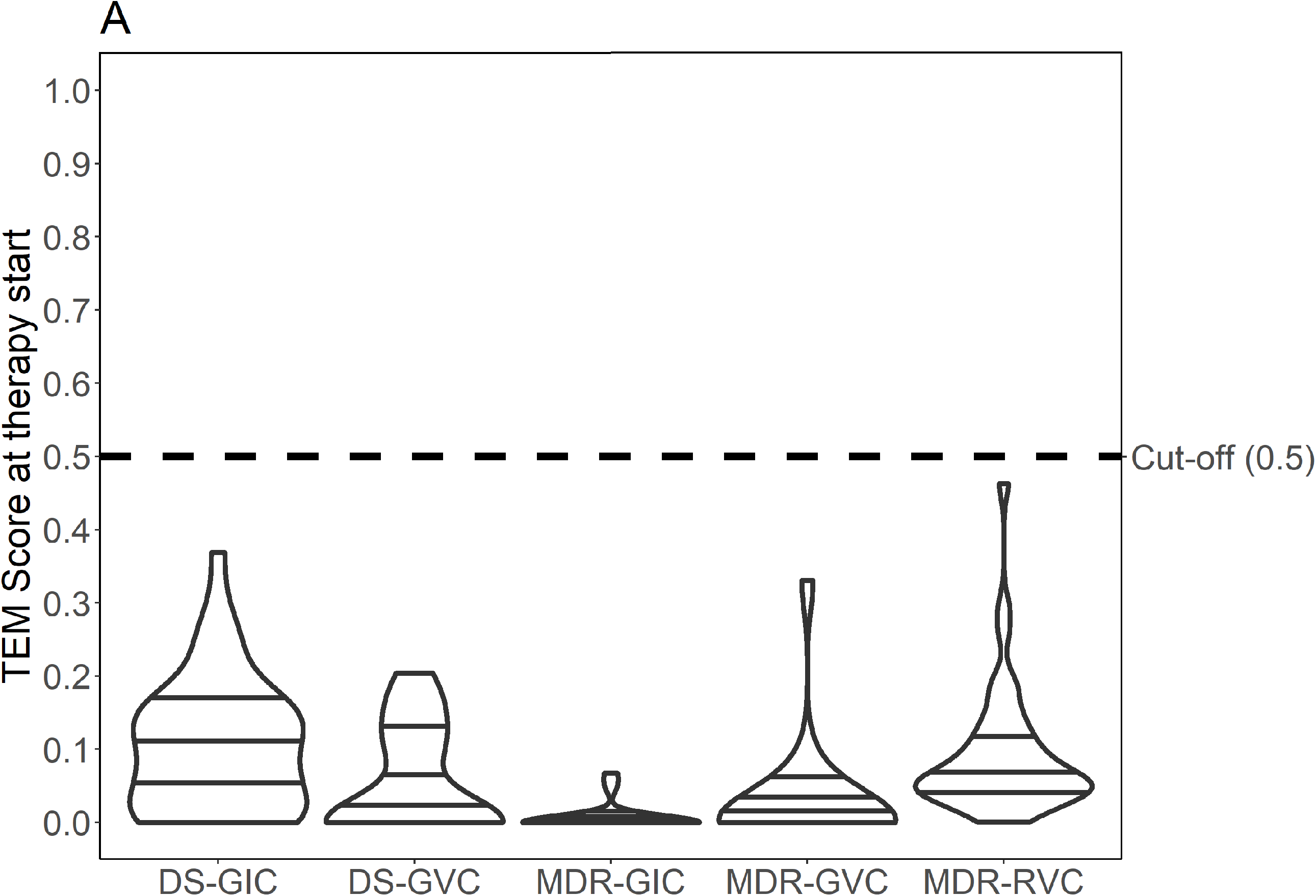
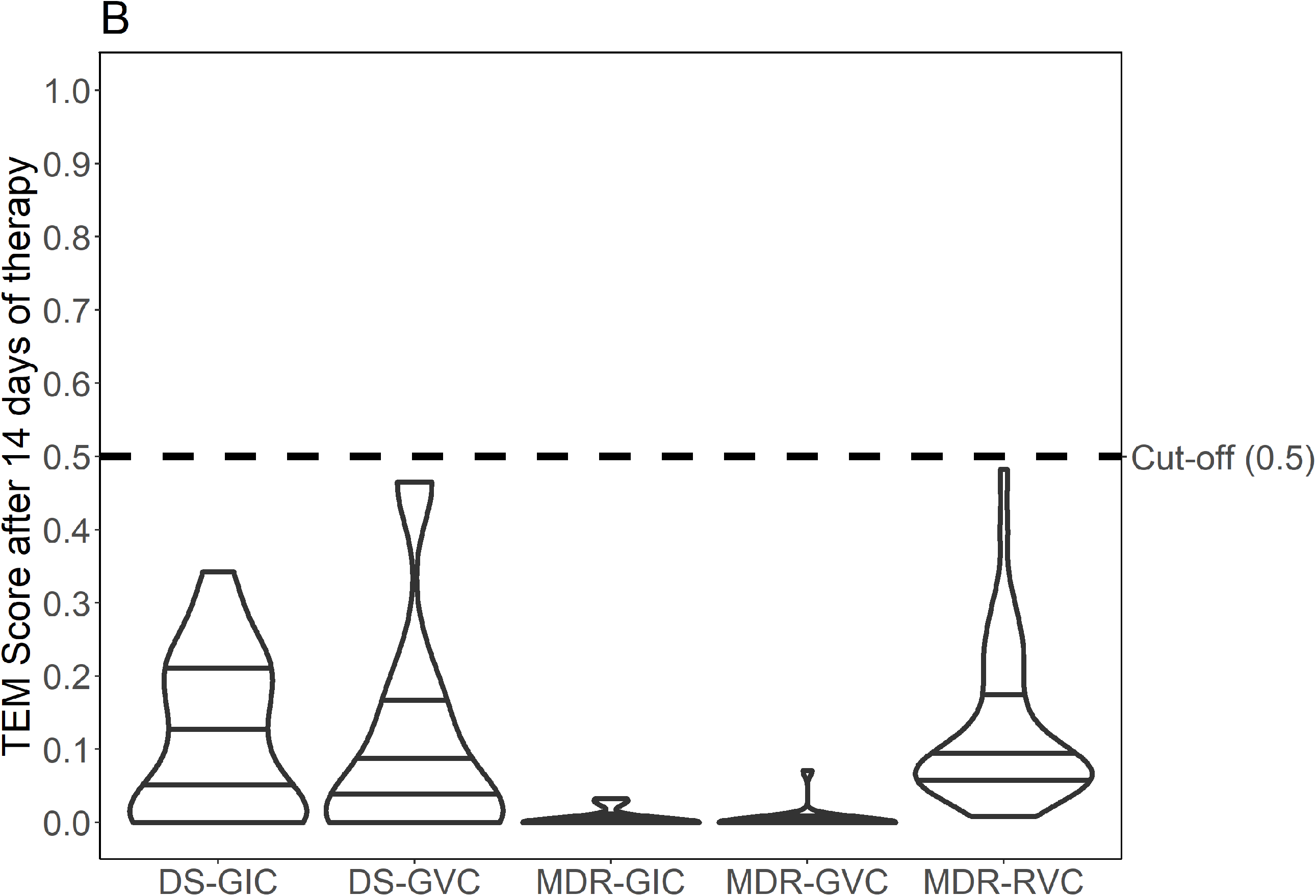
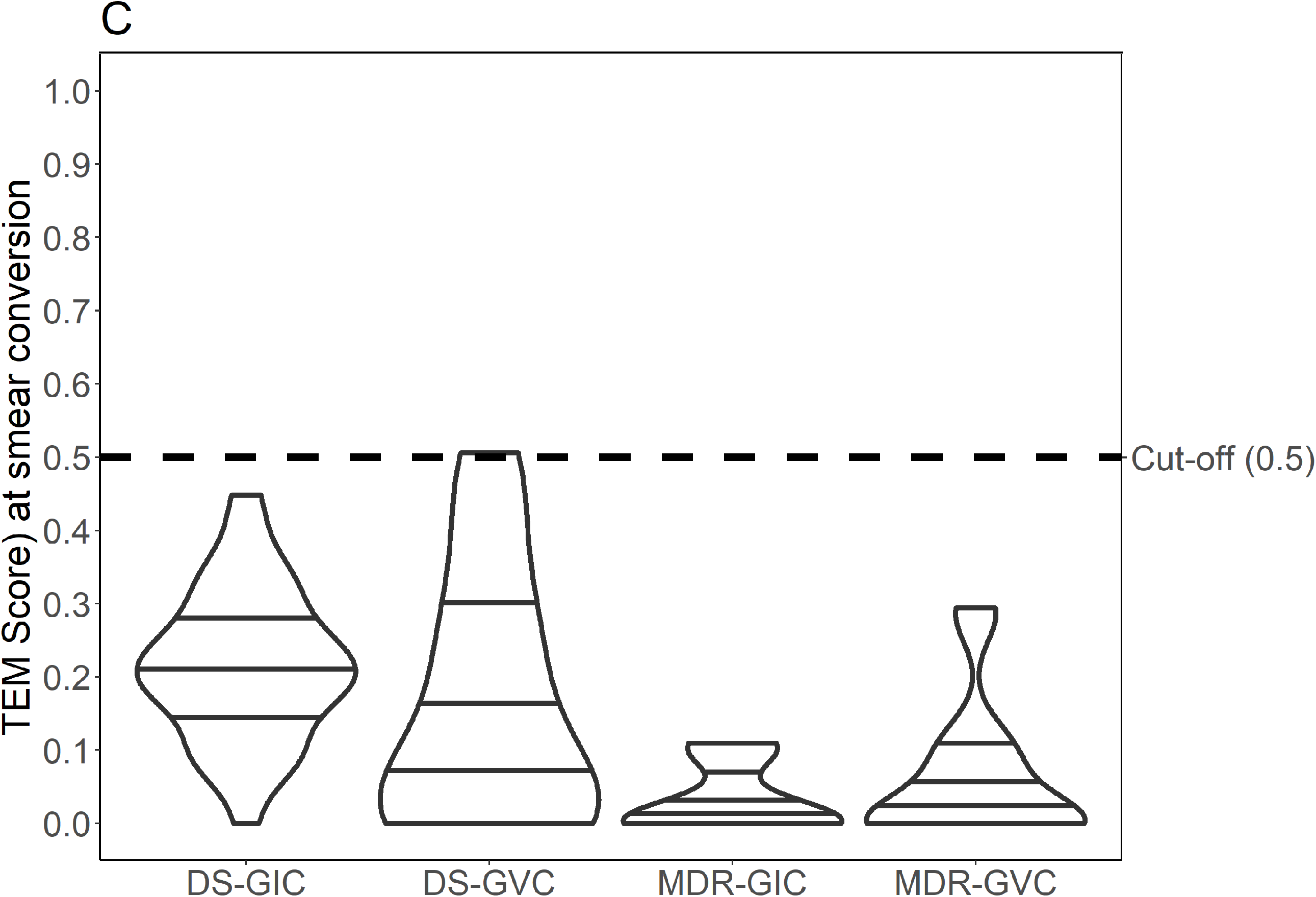
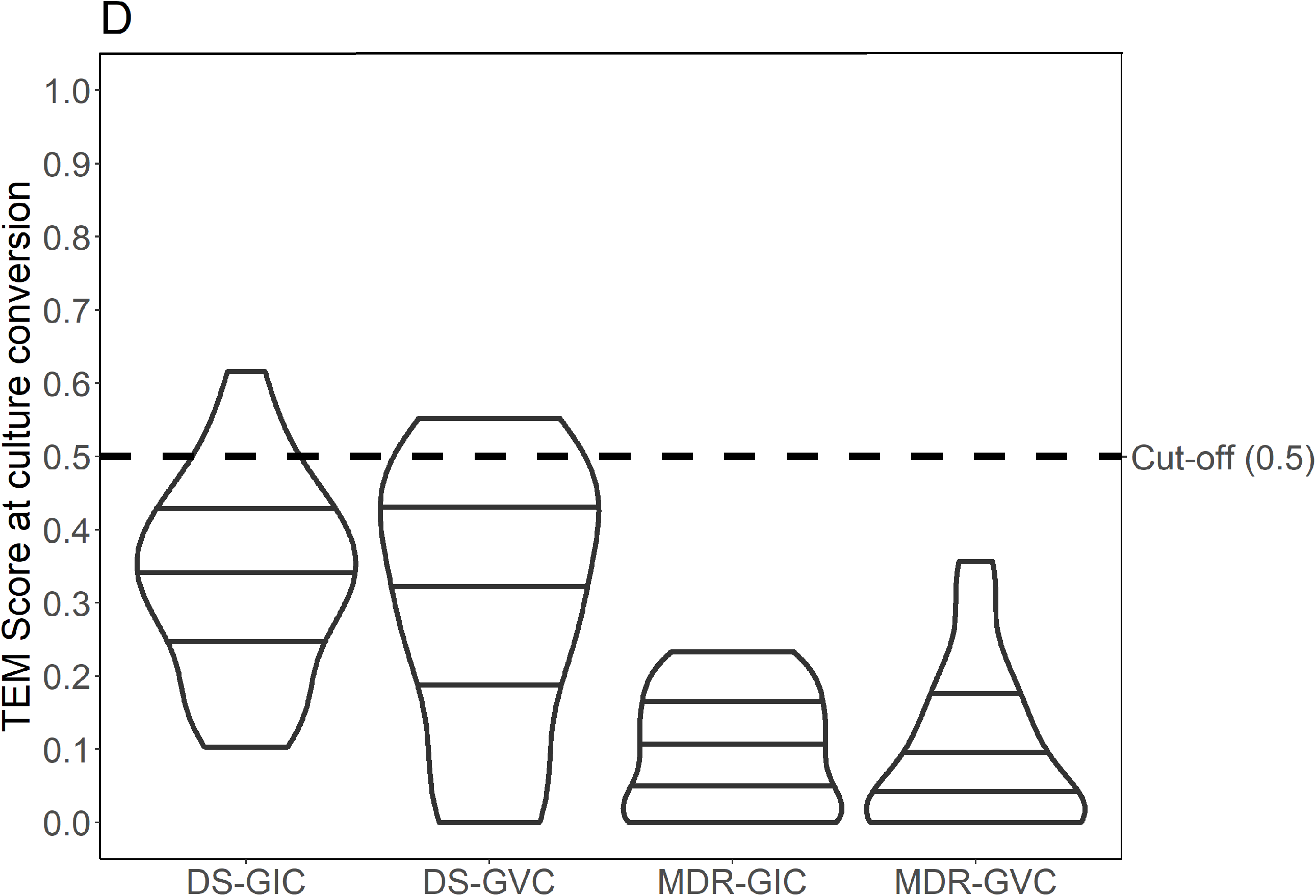
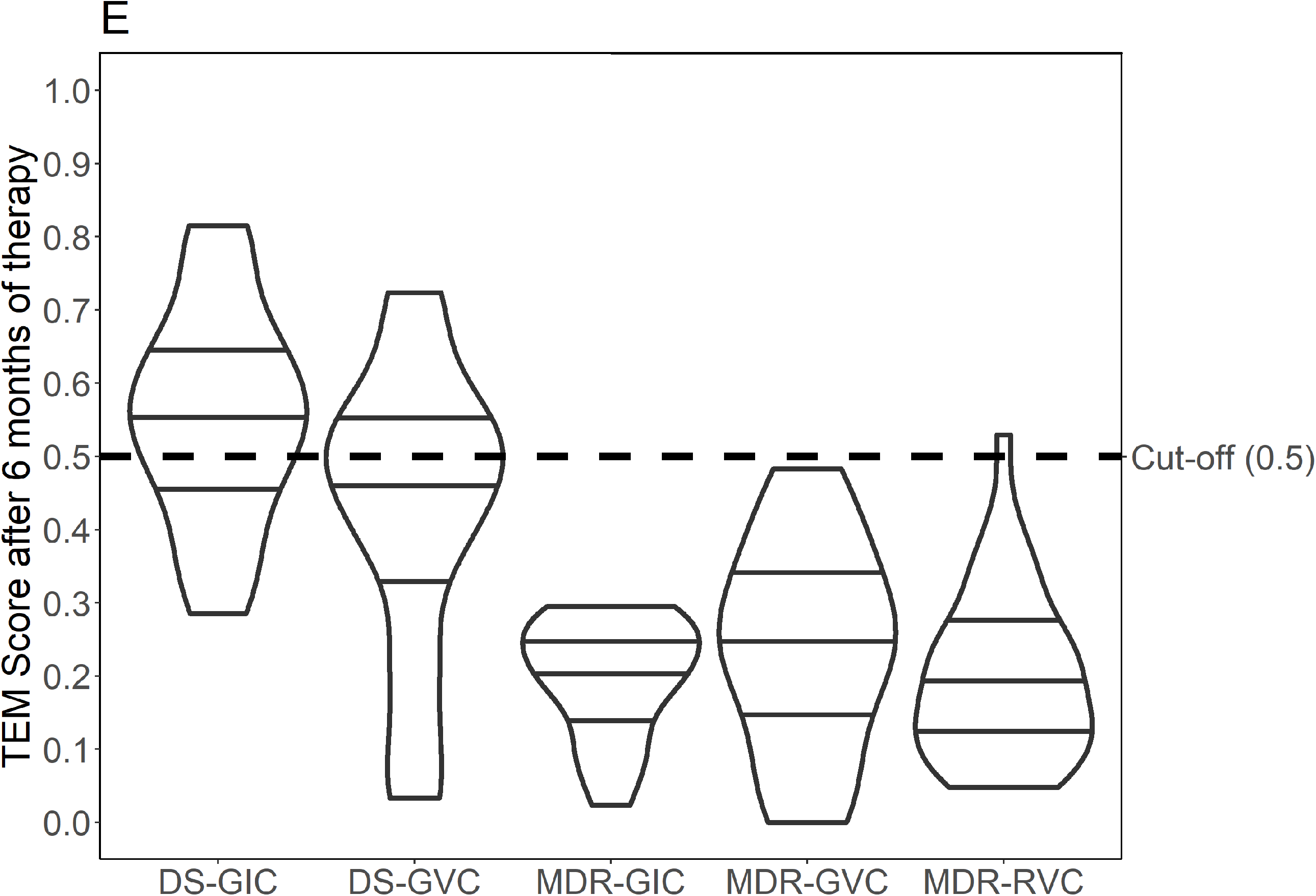
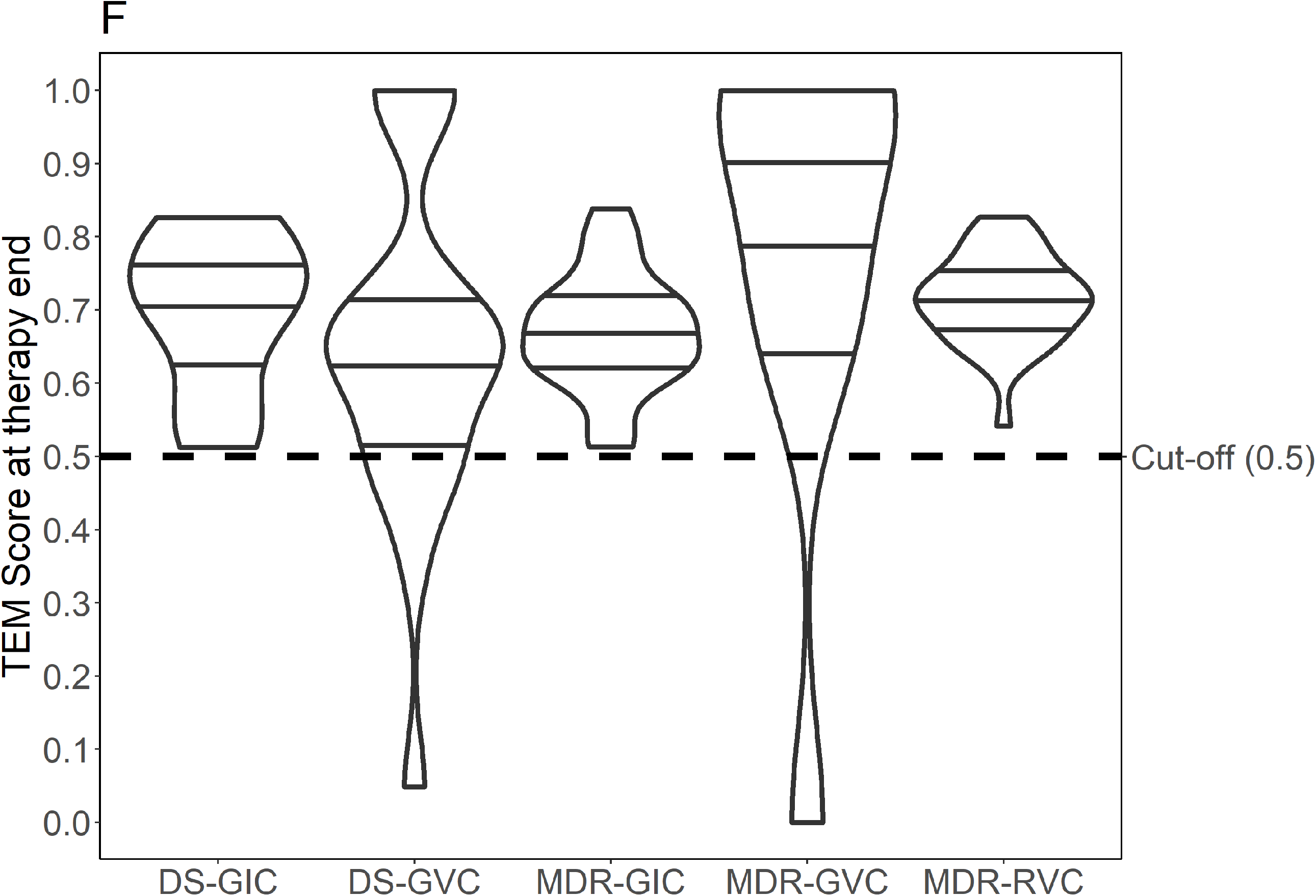
Therapy end model scores for the different cohorts of patients with tuberculosis at relevant clinical timepoints. Violin plot with therapy end model (TEM) scores for drug susceptible (DS) tuberculosis patients from the German identification cohort (DS-GIC) and the German validation cohort (DS-GVC), and for multidrug-resistant (MDR) tuberculosis patients from the GIC, the GVC, and the Romanian validation cohort (RVC) at different time points during therapy. Y-axis: TEM score, dotted horizontal line: threshold for relapse-free end-of-therapy (Scores (P)≥0.5); X-axis showing the different cohorts. **Figure 4A:** TEM scores at therapy start for DS-GIC, DS-GVC, MDR-GIC, MDR-GVC, and MDR-RVC patients, **Figure 4B:** TEM scores at 14 days of therapy for DSGIC, DS-GVC, MDR-GIC, MDR-GVC, and MDR-RVC patients, **Figure 4C:** TEM scores at smear conversion for DS-GIC, DS-GVC, MDR-GIC, and MDR-GVC patients, **Figure 4D:** TEM scores at culture conversion for DS-GIC, DS-GVC, MDR-GIC, and MDR-GVC patients, **Figure 4E:** TEM scores at 6 months of therapy for DS-GIC, DS-GVC, MDR-GIC, MDR-GVC, and RVC-MDR patients, and **Figure 4F:** TEM scores at individual therapy end for DS-GIC, DS-GVC, MDR-GIC, MDR-GVC, and RVC-MDR patients.

The calculated therapy durations did not differ significantly from observed durations for the DS-GIC patients (median calculated 175.0 days vs. observed 184.0 days, p=0.104) but they do for the DS-GVC group (median calculated 225 vs. observed 273.0 days, p=0.001), which could be explained by the larger gaps between sampling timepoints, or higher bacillary burden at baseline. Calculated therapy durations were significantly shorter compared to those observed in patients of the MDR-GIC (median calculated 420.0 days vs. observed 638 days, p=0.001) and the MDR-GVC group (median calculated 430.0 days vs. observed 641 days, p<0.001). Calculated therapy durations in MDR-RVC patients were also significantly shorter than the observed durations (median calculated 450.0 vs. observed 611.0 days, p=0.001). For patients in the MDR-GIC, this would have resulted in a median reduction of therapy duration by 218 days. In the MDR-GVC, therapy would have been reduced by a median of 211 days, and for a median of 161 days in the MDR-RVC.

## Discussion

Whole-blood-derived RNA transcriptomic analysis in samples from two cohorts of tuberculosis patients in Germany, one with drug-susceptible tuberculosis and one with multidrug-resistant tuberculosis, yielded a 22-gene RNA therapy end model that was able to indicate individual therapy durations associated with cure with high accuracy. The therapy end model was subsequently applied to two independent validation cohorts of patients with drug-susceptible- and multidrug-resistant tuberculosis from Germany and a third validation cohort of patients with multidrug-resistant tuberculosis from Romania. The therapy end model gave individual probabilities for cure-associated end-of-therapy time points at any given moment throughout therapy, which therefore provided data for therapy monitoring. Adjusting treatment durations based on the model’s probabilities could reduce the overall MDR-TB treatment time considerably.

Transcriptional signatures for the prediction of progression to active diseases, the diagnosis of tuberculosis, and early responses to anti-tuberculosis therapy in patients with drug-susceptible tuberculosis have been published (*12, 17, 18*). Clinical therapy outcomes, including recurrent disease, have also been predicted by published RNA signatures (*12, 13, 19*). In contrast to these studies, hereby described therapy end model was able to indicate individual therapy durations among patients with drug-susceptible- and multidrug-resistant tuberculosis. Compared to other published marker combinations, our findings were affirmed by involving various established clinical endpoints such as smear and culture status, radiological findings, and strict outcome criteria (*14, 16, 20*), which include a follow-up period of one year after completion of therapy to capture disease recurrence. Application of the model to patients with drug-susceptible tuberculosis was able to identify end-of-therapy timepoints with high accuracy. In addition, the model uniquely provides individualized therapy durations in patients with multidrug-resistant tuberculosis.

Therapy responses in patients with tuberculosis are usually evaluated by serial sputum culture sampling, which are not accessible during later stages of therapy in most cases. Therefore, culture is not an accurate marker to guide individualized therapy durations (*21*). Of note, the probability levels for cure classified by this model at defined biological timepoints, i.e. sputum smear or culture conversion, were comparable in patients with drug-susceptible- and multidrug-resistant tuberculosis. We conclude that the 22-gene RNA therapy end model is a promising biomarker to monitor treatment responses not only early in therapy but also at later stages of therapy when *M. tuberculosis* cultures are no longer available.

Our model yielded shorter treatment durations for most patients with multidrug-resistant tuberculosis enrolled in this study. There have been standardized approaches recommended for a shorter multidrug-resistant tuberculosis treatment regimen with therapy durations of 9–11 months (*22*). This regimen has been utilized in patients with multidrug-resistant tuberculosis globally leading to successful outcomes in a high proportion of study patients (*23*). However, *M. tuberculosis* isolates from European patients with multidrug-resistant tuberculosis frequently carry second-line drug-resistance against core drugs included in the shorter course multidrug-resistant tuberculosis regimen; hence, the regimen’s use among European patients is severely limited (*24*). Standardized therapies have been shown to lead to higher proportions of treatment failure and disease relapse in settings with high proportions of drug-resistances when compared to individualized therapy regimens (*25*). Therefore, tailored therapies informed by comprehensive drug-resistance testing may be a more promising approach to design effective multidrug-resistant tuberculosis drug regimens (*14, 26*). The therapy end model described herein can add substantial value to the individualized therapy approach since the drug regimens’ effect can be monitored and individual durations can be precisely identified. This approach has the potential to pave the way to individualized treatment regimens that are shorter and more effective than the WHO’s currently recommended, standardized treatment regimen (*22*).

Individualized durations largely depend on the bacterial load, the host constitution, the pathogen’s resistance pattern, and the availability of drugs. RNA signature-guided individualized therapy with shorter treatment duration can potentially avert disease recurrence, lessen adverse events, improve compliance and reduce overall cost for tuberculosis treatment, particularly among patients with multidrug-resistant tuberculosis. Future clinical evaluation of individualized therapy durations in patients with tuberculosis requires comparative studies such as a non-inferiority approach, which has demonstrated the general usefulness of shorter multidrug-resistant tuberculosis treatment regimens (*23*). Importantly, the full impact of RNA signature-guided individual therapies cannot be realized without the development of an affordable point-of-care assay and platform amenable to implementation in high burden, low-income countries.

In clinical practice, most patients treated for drug-susceptible tuberculosis using the standard four-drug regimen are cured before completing 6 months of therapy (*27*). This study was designed to identify and validate biomarkers for individualized therapy durations in patients with multidrug-resistant tuberculosis, not in patients with drug-susceptible disease. Therefore, the sampling schedule did not include fixed study visits between month 4 and 6 of anti-tuberculosis therapy to detect possible end-of-therapy timepoints in patients with drug-susceptible tuberculosis. A more frequent sampling strategy could have provided data to calculate more precise end-of-therapy timepoints for patients with drug-susceptible tuberculosis possibly indicating shorter durations of therapy.

The therapy end model was mainly based on the outcome definitions provided by the TBNET criteria (*14, 16, 20*). The model’s performance to discriminate between cure and failure in this study was limited to those patients who had a negative or positive *M. tuberculosis* culture status at 6 months of therapy (*16*) since we did not observe patients experiencing relapse within one year of post treatment follow-up. One of the advantages of the TBNET treatment outcome definitions, in contrast to WHO treatment outcome definitions, is that they evaluate the parameter “cure” one year after the end of therapy and thus consider also relapse within this period (*16*).

The study was mainly conducted in Caucasian patients and did not include people living with HIV, where RNA expression data analysis may yield different results. Still, patients from this prospective multi-center trial had an in-depth clinical and bacteriological observational follow-up schedule.

In conclusion, we prospectively identified and validated a host 22-gene RNA-based therapy end model to indicate individual end-of-treatment timepoints to achieve cure in patients treated for drug-susceptible- and multidrug-resistant tuberculosis. This model has the potential to significantly shorten treatment duration in the majority of patients with multidrug-resistant and potentially with drug-susceptible tuberculosis. The model’s translation into clinical practice will require further clinical evaluation and the development of an implementable platform to support feasibility in resource limited settings.

## Materials and Methods

### Study design and participants

Between March 2013 and March 2016, culture-confirmed patients with pulmonary drug-susceptible tuberculosis (including mono-or polydrug resistance, but excluding rifampicin-resistant tuberculosis), and multidrug-resistant tuberculosis identified by detection of *M. tuberculosis* DNA from sputum by the Xpert MTB/RIF test (Cepheid, Sunnyvale, USA) were prospectively enrolled into the DS-GIC and the MDR-GIC, respectively. The enrolling institutions were the Medical Clinic, Research Center Borstel; Karl-Hansen-Klinik, Bad Lippspringe; Sankt Katharinen-Krankenhaus, Frankfurt; Thoraxklinik-Heidelberg, Heidelberg; Asklepios Fachkliniken München-Gauting, Munich, all in Germany, as previously described (*14, 15*) Between March 2015 and April 2018, patients with drug-susceptible tuberculosis and multidrug-resistant tuberculosis were prospectively enrolled into the DS-GVC and MDR-GVC at the same centres and additionally at the Klinikum Dortmund, Germany, and at the University Clinic of Cologne, Germany. Between May 2015 and March 2017, patients with multidrug-resistant tuberculosis were prospectively enrolled into the MDR-RVC at the Marius-Nasta-Institute (MNI) in Bucharest, Romania. Individuals were not included to the study, if they were less than 18 years of age, under legal supervision, or living with human immunodeficiency virus (HIV).

Between June 2015 and December 2015, adult HCs with no history of previous tuberculosis and without any known concurrent illnesses at the timepoint of blood sampling were enrolled at the Medical Clinic of the Research Center Borstel (Germany).

Study visits included clinical assessment and blood sampling for whole-blood RNA measurements from PAXgene tubes (Qiagen®, Venlo, the Netherlands). Study visits were performed at ideally before treatment initiation, at 14 days of therapy, at the times of smear conversion and following culture conversion (not available in the MDR-RVC), at 6 months and/or therapy end in patients with drug-susceptible tuberculosis, and additionally, at 10, 15, 20 months of therapy in patients with multidrug-resistant tuberculosis. After completion of 4 weeks of therapy, an additional study visit was performed in patients from the MDR-RVC. All patients completed 12 months of evaluation following end-of-therapy to capture disease recurrence. A subset of DS/MDR-GVC participants also provided specimens during this follow-up period. Sputum samples provided by German study participants were evaluated via smear microscopy and culture at the National Reference Center for Mycobacteria at the Research Center Borstel. Samples provided by study participants at the MNI were analyzed at the Romanian National Reference Center for Mycobacteria in Bucharest. Anti-tuberculosis therapy regimens were based on comprehensive drug-susceptibility testing and consistent with current therapy recommendations (*5, 28, 29*). Treatment outcomes were assessed following the TBNET definitions, where relapse-free cure is defined by having a negative *M. tuberculosis* culture status at 6 months after treatment initiation without positive cultures thereafter and no disease recurrence during the follow-up period of one year after therapy end (*16*). Treatment failure is defined by at least one positive *M. tuberculosis* culture 6 months after treatment initiation or thereafter, or a relapse within one year after treatment completion (*16*). TBNET outcome criteria were preferred since the WHO outcome definitions do not include one-year follow-up post treatment completion to exclude for recurrent disease (**Table 3**). In addition, the WHO’s definition for treatment success involves certain items that cannot be predicted by a biomarker since they depend on the patient’s behavior or clinical decisions in the course of therapy (i.e. treatment completion or change of drugs during the course of treatment) (*30*).

**Table 3:** Treatment outcome definitions in multidrug-resistant tuberculosis according to WHO 2014 (*30*) and TBNET simplified definitions 2016 (*16*). n/a: not applicable.

|  | WHO Definition - revised 2014 | Simplified definitions (TBNET) |
| --- | --- | --- |
| Cured | Treatment completed as recommended by the national policy without evidence of failure AND three or more consecutive cultures taken at least 30 days apart are negative after the intensive phase | A negative culture status 6 months after treatment initiation, no positive cultures thereafter, and no relapses within 1 year after treatment completion |
| Treatment completed | Treatment completed as recommended by the national policy without evidence of failure BUT no record that three or more consecutive cultures taken at least 30 days apart are negative after the intensive phase. | n/a |
| Treatment failed | Treatment terminated or need for permanent regimen change of at least two anti-TB drugs because of:<br>– lack of conversion by the end of the intensive phase, or<br>– bacteriological reversion in the continuation phase after conversion to negative, or<br>– evidence of additional acquired resistance to fluoroquinolones or second-line injectable drugs, or<br>– adverse drug reactions (ADRs). | A positive culture status 6 month after treatment initiation or thereafter, or a relapse within 1 year after treatment completion. |
| Died | A patient who dies for any reason during the course of treatment | A patient who dies for any reason during the course of observation |
| Lost to follow-up | A patient whose treatment was interrupted for 2 consecutive months or more | Non-receipt of care 6 months after treatment initiation. |
| Not evaluated | A patient for whom no treatment outcome is assigned. (This includes cases “transferred out” to another treatment unit and whose treatment outcome is unknown) | n/a |
| Undeclared | n/a | - no culture status at 6 months while the patient was receiving care, or<br>- no post-treatment assessment |
| Treatment success | The sum of cured and treatment completed | n/a |

### RNA processing, and data analysis

Whole blood RNA isolation from PAXgene (Qiagen®, Venlo, the Netherlands) was handled according to the manufacturer’s instructions and stored at −80°C until RNA isolation using the PAXgene blood RNA isolation kit (Qiagen®, Venlo, the Netherlands). Aliquots of isolated RNA were used for quality control to analyze the RNA integrity with the RNA Nano 6000 Kit on an Agilent Bioanalyzer (Agilent®, Böblingen, Germany) according to the manufacturer’s instructions. In case of an insufficient RNA Integrity Number (RIN) as a measure of sample quality and number or signs of degradation, samples were excluded from further analysis.

#### Labelling, hybridization and scanning of microarrays

Total RNA was used for reverse transcription and subsequent Cy3-labelling with the Low-Input Quick Amp Labeling Kit (Agilent®, Böblingen, Germany) according to the One-Color Microarray-Based Gene Expression Analysis Protocol version 6.9.1 (Agilent®, Böblingen, Germany) with RNA Spike-In controls. 1650ng of Cy3-labelled cRNA was hybridized to human 4x44K V2 gene expression microarrays according to manufacturer’s instructions. Arrays were scanned on a SureScan microarray scanner (Agilent®, Böblingen, Germany) at a resolution of 5 μm.

#### Data extraction and normalization

Raw expression data from scanned microarray slides were extracted from tiff files using the Feature Extraction Software version 11 (Agilent®, Böblingen, Germany). Raw data files were imported into Agilent GeneSpring software version 13 (Agilent®, Böblingen, Germany). Percentile Shift was used as normalization method with the 75th percentile as a target and baseline transformation was applied to the median of all samples. Prior to data analysis, quality control was performed and compromised probes removed from further analysis. “Normalized expression” will refer in the manuscript to Log2 transformed expression values.

#### Data analysis

Comparisons between cohorts were performed by Kruskal-Wallis Test. The data analyses were performed with the software R (versions 3.4.2 to 4.00). The general transcriptomic data preparation was performed with the packages limma, reshape2, and tidyr. The package ggplot2 was used for graphical data presentation. Therapy outcome estimations and the transformation of this factor to a numerical score for the development of the therapy outcome score were performed using the R packages caTools, tidyr, reshape2, randomForest, dplyr and pROC. The development of the therapy progress score was executed by using the R packages MASS, tidyr, reshape2, dplyr, glmnet and caTools. All available time points were included for the therapy progression score and the therapy end model regardless of therapy outcome status. The R packages car, caTools, tidyr, dplyr, glmnet, randomForest and pROC were used to develop the therapy end model. In general, the outcome “lost to follow-up” was not an exclusion criterion to calculate hypothetical end-of-therapy scores.

### Data availability

Microarray data have been deposited at Gene Expression Omnibus database (GSE147690, GSE147689, GSE147691).

### Ethics

Study approval was granted by the Ethics Committee of the University of Lübeck, Germany (AZ 12–233), which was then approved by the corresponding local Ethic Committees of all participating centers in Germany, and by the Ethics Committee of the Marius Nasta Institute (3181/25.03.2015; Bucharest, Romania).

## Supplementary Materials

**Supplementary File: List of genes of the different steps contributing to the therapy end model**.

The first Excel file tab shows the preselection of genes contributing to the therapy outcome score (TOS), the second tab the ones contributing to the therapy progression score (TPS), and the third one shows the genes identifies for the end-of-therapy (EOT) list. The TOS tab lists the identified differentially expressed genes of healthy controls when compared to drug-susceptible (DS) and multidrug-resistant (MDR) tuberculosis patients of the German Identification Cohort (GIC) at therapy start. The TPS tab shows genes that correlate with the remaining days of therapy of DS-and MDR-GIC patients as basis for the therapy progression score. The EOT tab shows genes that are differentially expressed in all time points of drug-susceptible GIC patients under anti-tuberculosis therapy when compared to the end-of-therapy as basis end-of-therapy list.

## Acknowledgments

We thank Jessica Hofmeister, Franziska Daduna, Sandra Nyenhuis, Frauke Koops, Lasse Möller, and Jasmin Tiebach for laboratory work. We thank Cordula Ehlers, Nelleke Smitsman, and Susanne Dox for study and data management.

## Author contributions

JH and CL made a substantial contribution to the conception and design of the work and to the acquisition of data. JH, MR, SM, TG, and CL made substantial contributions for the analysis and interpretation of data for the work, wrote the manuscript, critically revised the manuscript for important intellectual content, and gave final approval of the current version to be published. JH, MR and SM performed statistical analysis. KA, AD, GG, MH, EI, BK, SHEK, IK, FvL, AMM, FM, MM, DN, IDO, CP, AR, TR, JR, HJFS, PS-C, MS, DS, VS, IS, ET, MU, JW made contributions to the acquisition of the data for the work, critically revised the manuscript for important intellectual content, and gave final approval of the current version to be published. All authors agree to be accountable for all aspects of the work in ensuring that questions related to the accuracy or integrity of any part of the work are appropriately investigated and resolved.

